# Effectiveness of BNT162b2 mRNA vaccine third doses and previous infection in protecting against SARS-CoV-2 infections during the Delta and Omicron variant waves; the UK SIREN cohort study September 2021 to February 2022

**DOI:** 10.1101/2023.05.22.23290197

**Authors:** Victoria J Hall, Ferdinando Insalata, Sarah Foulkes, Peter Kirwan, Dominic Sparkes, Ana Atti, Michelle Cole, Elen de Lacy, Lesley Price, Diane Corrigan, Colin S Brown, Jasmin Islam, The SIREN Study Group Andre Charlett, Andre Charlett, Susan Hopkins

## Abstract

Third doses of COVID-19 vaccines were widely deployed following primary vaccine course waning and emergence of the Omicron-variant. We investigated protection from third-dose vaccines and previous infection against SARS-CoV-2 infection during Delta-variant and Omicron-variant (BA.1 & BA.2) waves in our frequently PCR-tested cohort of healthcare-workers.

---

Relative effectiveness of BNT162b2 third doses and infection-acquired immunity was assessed by comparing the time to PCR-confirmed infection in boosted participants with those with waned dose-2 protection (≥254 days after dose-2). Follow-up time was divided by dominant circulating variant: Delta 07 September 2021 to 30 November 2021, Omicron 13 December 2021 to 28 February 2022. We used a Cox regression model with adjustment/stratification for demographic characteristics and staff-type. We explored protection associated with vaccination, infection and both.

We included 19,614 participants, 29% previously infected. There were 278 primary infections (4 per 10,000 person-days of follow-up) and 85 reinfections (0.8/10,000 person-days) during the Delta period and 2467 primary infections (43/10,000 person-days) and 881 reinfections (33/10,000) during the Omicron period. Relative Vaccine Effectiveness (VE) 0-2 months post-3^rd^ dose (V3) (3-doses BNT162b2) in the previously uninfected cohort against Delta infections was 63% (95% Confidence Interval (CI) 40%-77%) and was lower (35%) against Omicron infection (95% CI 21%-47%). For primary course ChAdOX1 recipients, BNT162b2 heterologous third doses were especially effective, with VE 0-2 months post-V3 over ≥68% higher for both variants. Third-dose protection waned rapidly against Omicron, with no significant difference between two and three BNT162b2 doses observed after 4- months. Previous infection continued to provide additional protection against Omicron (67% (CI 56%-75%) 3-6 months post-infection), but this waned to about 25% after 9-months, approximately three times lower than against Delta.

Infection rates surged with Omicron emergence. Third doses of BNT162b2 vaccine provided short-term protection, with rapid waning against Omicron infections. Protection associated with infections incurred before Omicron was markedly diminished against the Omicron wave. Our findings demonstrate the complexity of an evolving pandemic with potential emergence of immune-escape variants and the importance of continued monitoring.

## BACKGROUND

Third doses of COVID-19 vaccines (V3) were widely deployed in winter 2021/2022 in the United Kingdom (UK) and other high-income countries (source our world in data). Third doses were first deployed in the Autumn 2021 (Delta-variant dominant period), targeted in the UK to those considered at higher risk of severe disease (older age, immunosuppressed) and to those providing healthcare to these individuals,^1^ in response to mounting evidence of waning vaccine effectiveness against symptomatic illness,^2–5^ and infection,^6–8^ six months after administration of the second dose. Neutralising antibody titres, associated with vaccine efficacy, were shown to be greatly increased by a booster vaccination.^9–12^

In late November 2021, with the rapid emergence and rapid spread of the Omicron variant, access to third doses was widened to all those over the age of 18 years as a response measure in the UK.^13^ With over five-fold more mutations in the Spike and Receptor Binding Domain (RBD) proteins compared to the Delta-variant, higher rates of evasion from both vaccine-and infection-acquired immunity were anticipated.^14, 15^

The SARS-CoV-2 Immunity and Reinfection Evaluation (SIREN) study, a large prospective cohort study of UK healthcare workers undergoing fortnightly PCR screening, has tracked primary infections and reinfections continuously since June 2020. Conducting daily monitoring to identify reinfections and vaccine breakthroughs, Omicron was rapidly identified as a step-change in our cohort, with rates of reinfections rapidly rising in the cohort, despite vaccination.

In this analysis we investigate the effectiveness of third doses with BNT162b2 vaccines, including the impact of hybrid-immunity against SARS-CoV-2 infection in our frequently PCR-tested cohort of healthcare-workers.

## METHODS

### Study Design and Oversight

The SIREN study is an ongoing, multicentre, prospective cohort study involving health care workers (≥18 years of age) in the United Kingdom (ISRCTN 11041050)^16^. All the authors vouch for the accuracy and completeness of the data and for the fidelity of the study to the protocol.

### Participant Involvement

At its core the SIREN study is a collaboration between participants and researchers. We are committed to ensuring participant feedback is heard, respected, and influences our research. The SIREN study runs a comprehensive engagement programme providing participants with regular opportunities to engage with, and feedback on, our research. The SIREN Participant Involvement Panel (PIP) is central to this, providing timely ongoing participant input into cohort recruitment and retention, study developments and communications. PIP members represent the diversity of the SIREN cohort and are supported by experts in participant engagement research.

### Study Participants and Data

Participants underwent PCR testing for SARS-CoV-2 every two weeks, supplemented by widespread lateral-flow testing, and regular (monthly or quarterly) antibody testing. Data sources have been described previously.^6^ Participants were included in this analysis if they had received at least two doses of COVID-19 vaccine and their previous infection status at the point of entry to the analysis period was known. Participants were excluded if the outcome could not be determined (e.g., if they did not undergo PCR testing during the follow-up period), or if the date of onset of the primary infection, based on either a positive PCR test or COVID-19 symptoms, was not available. We included participants whose primary vaccine course was either BNT162b2 vaccine (Pfizer–BioNTech) or ChAdOx1 nCoV-19 vaccine (AstraZeneca), and whose booster vaccine (if received) was BNT162b2 vaccine (Pfizer–BioNTech). Participants with other vaccine combinations were excluded because of the small numbers involved.

### Primary Outcome

The primary outcome was a PCR-confirmed SARS-CoV-2 infection, irrespective of the participant’s symptom status. This outcome was defined as a primary infection in the previously uninfected cohort or a reinfection in the previously infected cohort (two PCR-positive samples ≥90 days apart or, if the primary infection PCR is not available, a new PCR-positive sample ≥28 days after an antibody-positive result consistent with previous infection).

Participants could move from the previously uninfected cohort to the previously infected cohort 90 days after their PCR-positive test. For individual participants, the end of follow-up was the date of primary infection (in the previously uninfected cohort), the date of reinfection (in the previously infected cohort), or the date of the last PCR-negative test.

### Statistical Analysis

In our Cox proportional-hazards model, the outcome was time to PCR-positive SARS-CoV-2 infection, and main predictors — vaccination status and previous infection status — were categorical and time-varying. We stratified according to age group, geographic region, and frequency of exposure to persons with Covid-19. We chose stratification based on these categorical predictors because they were statistically significant when controlled for but did not satisfy the proportional-hazards assumption (Schoenfeld test, according to predictor and global fit). We also controlled for sex, ethnic group and workplace setting (see Supplementary tables A.1 through A.6), because we observed that these predictors were statistically significant, led to a significant increase in the likelihood value (likelihood ratio test) and satisfied the proportional-hazards assumptions In Tables 1-3, we also report estimates from an unadjusted model, without stratifying or controlling for any predictor other than the time since vaccination and infection. The model accounted for calendar time, which is crucial given the varying infection rate, through the baseline hazard that can take any functional form. In this model, the hazard is assumed to be

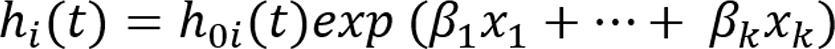

with a time-varying baseline hazard 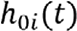 for each stratum. We estimated the parameter β, report the hazard ratio HR=exp(β), and report vaccine effectiveness and protection from primary infection calculated as 1 minus the hazard ratio, along with Wald statistic confidence intervals. The estimates of the hazard ratios are independent of the baseline hazard, on which no assumption was made.

**Table 1:**
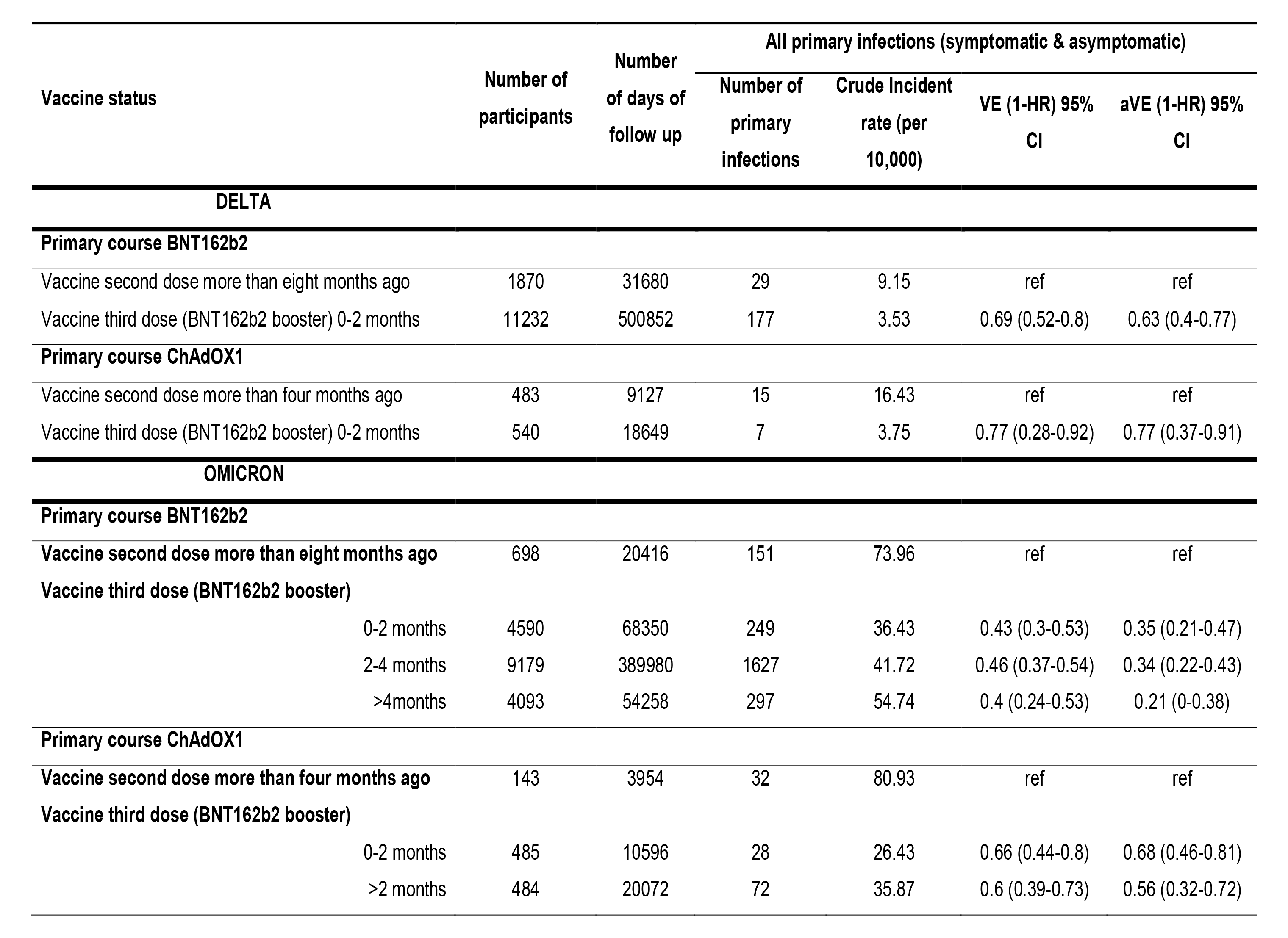
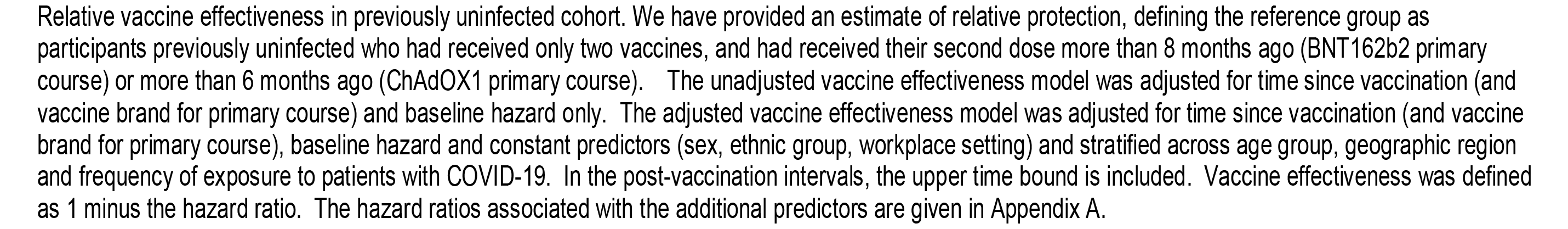
Vaccine effectiveness in the previously uninfected cohort against Delta and Omicron infections, September 2021 – February 2022.

| Vaccine status | Number of participants | Number of days of follow up | All primary infections (symptomatic & asymptomatic) |  |  |  |
| --- | --- | --- | --- | --- | --- | --- |
|  |  |  | Number of primary infections | Crude Incident rate (per 10,000) | VE (1-HR) 95% CI | aVE (1-HR) 95% CI |
| DELTA |  |  |  |  |  |  |
| Primary course BNT162b2 |  |  |  |  |  |  |
| Vaccine second dose more than eight months ago | 1870 | 31680 | 29 | 9.15 | ref | ref |
| Vaccine third dose (BNT162b2 booster) 0-2 months | 11232 | 500852 | 177 | 3.53 | 0.69 (0.52-0.8) | 0.63 (0.4-0.77) |
| Primary course ChAdOX1 |  |  |  |  |  |  |
| Vaccine second dose more than four months ago | 483 | 9127 | 15 | 16.43 | ref | ref |
| Vaccine third dose (BNT162b2 booster) 0-2 months | 540 | 18649 | 7 | 3.75 | 0.77 (0.28-0.92) | 0.77 (0.37-0.91) |
| OMICRON |  |  |  |  |  |  |
| Primary course BNT162b2 |  |  |  |  |  |  |
| Vaccine second dose more than eight months ago | 698 | 20416 | 151 | 73.96 | ref | ref |
| Vaccine third dose (BNT162b2 booster) |  |  |  |  |  |  |
| 0-2 months | 4590 | 68350 | 249 | 36.43 | 0.43 (0.3-0.53) | 0.35 (0.21-0.47) |
| 2-4 months | 9179 | 389980 | 1627 | 41.72 | 0.46 (0.37-0.54) | 0.34 (0.22-0.43) |
| >4months | 4093 | 54258 | 297 | 54.74 | 0.4 (0.24-0.53) | 0.21 (0-0.38) |
| Primary course ChAdOX1 |  |  |  |  |  |  |
| Vaccine second dose more than four months ago | 143 | 3954 | 32 | 80.93 | ref | ref |
| Vaccine third dose (BNT162b2 booster) |  |  |  |  |  |  |
| 0-2 months | 485 | 10596 | 28 | 26.43 | 0.66 (0.44-0.8) | 0.68 (0.46-0.81) |
| >2 months | 484 | 20072 | 72 | 35.87 | 0.6 (0.39-0.73) | 0.56 (0.32-0.72) |

Relative vaccine effectiveness in previously uninfected cohort. We have provided an estimate of relative protection, defining the reference group as participants previously uninfected who had received only two vaccines, and had received their second dose more than 8 months ago (BNT162b2 primary course) or more than 6 months ago (ChAdOX1 primary course). The unadjusted vaccine effectiveness model was adjusted for time since vaccination (and vaccine brand for primary course) and baseline hazard only. The adjusted vaccine effectiveness model was adjusted for time since vaccination (and vaccine brand for primary course), baseline hazard and constant predictors (sex, ethnic group, workplace setting) and stratified across age group, geographic region and frequency of exposure to patients with COVID-19. In the post-vaccination intervals, the upper time bound is included. Vaccine effectiveness was defined as 1 minus the hazard ratio. The hazard ratios associated with the additional predictors are given in Appendix A.

**Table 2:** Protection from infection over time, controlled for vaccine status, during Delta-variant and Omicron-variant periods.

| Vaccine status | Number of participants | Number of days of follow up | All reinfections (symptomatic & asymptomatic) |  |  |  |
| --- | --- | --- | --- | --- | --- | --- |
|  |  |  | Number of (re)infections | Crude Incident rate (per 10,000) | Protection (1-HR) 95% CI | Adjusted Protection (1-HR) 95% CI |
| DELTA |  |  |  |  |  |  |
| No previous infection | 19103 | 1935033 | 1125 | 5.81 | Ref | ref |
| Previous infection |  |  |  |  |  |  |
| 3-6 months | 718 | 19590 | 1 | 0.51 | 0.90 (0.39-0.98) | 0.90 (0.42-0.98) |
| 6-9 months | 2006 | 89006 | 13 | 1.46 | 0.77 (0.61-0.86) | 0.75 (0.59-0.85) |
| 9-12 months | 2515 | 143138 | 34 | 2.38 | 0.61 (0.44-0.72) | 0.63 (0.48-0.74) |
| 12-15 months | 1953 | 59925 | 2 | 0.33 | 0.93 (0.71-0.98) | 0.93 (0.72-0.98) |
| More than 15 months | 4595 | 398639 | 35 | 0.88 | 0.85 (0.81-0.89) | 0.85 (0.81-0.89) |
| OMICRON |  |  |  |  |  |  |
| No previous infection | 11074 | 569635 | 2467 | 43.31 | ref | ref |
| Previous infection |  |  |  |  |  |  |
| 3-6 months | 784 | 30397 | 47 | 15.46 | 0.66 (0.55-0.74) | 0.67 (0.56-0.75) |
| 6-9 months | 243 | 5855 | 14 | 23.91 | 0.50 (0.11-0.72) | 0.53 (0.2-0.73) |
| 9-12 months | 970 | 23871 | 80 | 33.51 | 0.19 (-0.07-0.38) | 0.27 (0.04-0.44) |
| 12-15 months | 1660 | 61798 | 214 | 34.63 | 0.22 (0.1-0.33) | 0.27 (0.16-0.37) |
| More than 15 months | 3099 | 150276 | 526 | 35.00 | 0.20 (0.1-0.29) | 0.26 (0.17-0.34) |

Protection from reinfection associated with a primary infection (PI). We have controlled for vaccine status without interaction terms between previous infection and vaccine status, after having verified that interaction terms are not significant (Wald tests: $p = 0.71$ for Delta period, $p = 0.67$ for Omicron period). The adjusted model includes additional predictors (gender, ethnic group, workplace setting) and is stratified across age group, geographic region and frequency of exposure to patients with COVID-19. In the post-infection intervals, the upper time bound is included. Protection is defined as 1 minus the hazard ratio. The hazard ratios associated with the additional predictors and the interaction models are given in Appendix A.

**Table 3:** Protection from infection and booster vaccination (hybrid protection) during Delta-variant and Omicron-variant periods

| Vaccine status | Number of participants | Number of days of follow up | All reinfections (symptomatic & asymptomatic) |  |  |  |
| --- | --- | --- | --- | --- | --- | --- |
|  |  |  | Number of reinfections | Crude Incident rate (per 10,000) | VE (1-HR) 95% CI | aVE (1-HR) 95% CI |
| DELTA |  |  |  |  |  |  |
| Vaccine two doses more than eight months ago and no previous infection | 1870 | 31680 | 29 | 9.15 | ref | ref |
| Vaccine three doses and infection less than one year ago – 0-2 months | 1715 | 55523 | 4 | 0.72 | 0.94 (0.81-0.98) | 0.88 (0.66-0.96) |
| Vaccine three doses and infection more than one year ago – 0-2 months | 2760 | 16249 | 2 | 1.23 | 0.94 (0.88-0.97) | 0.9 (0.76-0.96) |
| OMICRON |  |  |  |  |  |  |
| Vaccine two doses more than eight months ago and no previous infection | 698 | 20416 | 151 | 73.96 | ref | ref |
| Vaccine three doses and infection less than one year ago |  |  |  |  |  |  |
| 0-2 months | 901 | 14825 | 31 | 20.91 | 0.7 (0.56-0.79) | 0.66 (0.49-0.78) |
| 2-4 months | 1080 | 31993 | 70 | 21.88 | 0.72 (0.63-0.79) | 0.68 (0.57-0.76) |
| >4 months | 232 | 2698 | 6 | 22.24 | 0.76 (0.43-0.9) | 0.77 (0.33-0.92) |
| Vaccine three doses and infection more than one year ago |  |  |  |  |  |  |
| 0-2 months | 1909 | 34166 | 94 | 27.51 | 0.6 (0.48-0.69) | 0.57 (0.45-0.66) |
| 2-4 months | 3273 | 128921 | 457 | 35.45 | 0.54 (0.45-0.62) | 0.48 (0.38-0.57) |
| >4 months | 1313 | 16579 | 69 | 41.62 | 0.53 (0.37-0.66) | 0.41 (0.17-0.58) |

Protection from infection and booster vaccination (hybrid protection) during Delta-variant and Omicron-variant periods, in BNT162b2 recipients. We have provided an estimate of hybrid protection relative to a waned primary course, defining the reference group as participants previously uninfected who had received only two Pfizer vaccines, with second dose more than 8 months ago. In the post-vaccination intervals, the upper time bound is included. Protection is defined as 1 minus the hazard ratio. The adjusted vaccine effectiveness (aVE) model includes additional predictors (gender, ethnic group, workplace setting) and is stratified across age group, geographic region and frequency of exposure to patients with COVID-19. The hazard ratios associated with the additional predictors are given in Appendix A.

We conducted analyses separately for the periods of Delta-variant and Omicron-variant dominance, splitting follow-up time: Delta 07/09/2021 to 30/11/2021; Omicron 13/12/2021 to 28/02/2022. We defined three post-V3 time intervals for the Omicron period: 0 - 2 months, 2 - 4 months (upper bound included) and >4 months. For the Delta period we have a single interval approximately corresponding to the first two months after the third dose, before Omicron’s dominance. The baseline was either 8+ months after a second dose of BNT162b2, or, alternatively, 6+ months after a second dose of ChAdOx1. We first fitted the model in the previously uninfected cohort with vaccination status as a predictor with postvaccination (post-booster) intervals further categorized according to manufacturer of the vaccine administered in the primary course. The VE estimates represent the additional protection associated with a booster, relative to a waned primary course

We then performed analyses on both previously uninfected and previously infected participants, introducing previous infection status. First, we estimated the protection associated with primary infection. We grouped previous infection status into 6 categories: no primary infection, 3-6 months, 6<9, 9<12, 12-15 and >15 months after primary infection. We fitted a model where time since primary infection and time since vaccination are added as marginal predictors, without interactions. For the Delta period, we considered additional vaccine status categories (see table 2) in order to have enough follow-up time to obtain estimates. With this model we estimate protection associated with a previous infection, assuming that it is independent from vaccine status. In order to test this assumption, we fitted a model with interaction between the two predictors and verified that interaction terms were not significant. For this interaction model we split time since primary infection in only two categories, 3-12 and more than 12 months ago, to ensure convergence of hazard ratios on all combinations. We assumed that if interactions were not significant on a coarse scale, they would not appear in a more granular one. In Table 2 we only report the estimates for the previous infection categories, but not for vaccine status (see supplementary tables A.3 and A.4 for all estimates)

Next, we focussed on protection associated with third-dose vaccination and previous infection (hybrid immunity). We selected the recipients of the BNT162b2 vaccine and fitted a model with combination of the time since the primary infection (3-12 and more than 12 months ago) and the time since vaccination (table 3). The baseline here was a waned primary course of vaccination without any previous infection. The VE estimates from this model represent the combined effect associated with booster and previous infection. Recipients of the ChAdOx1 nCoV-19 vaccine for their primary course were excluded because of small numbers in the previously infected cohort.

Goodness of fit was assessed with the use of the likelihood ratio test (against the null model) and Akaike information values. The widths of the confidence intervals have not been adjusted for multiplicity and cannot be used to infer effects. We used robust variance estimates to guard against the potential for unmeasured confounders at the study site level.

We performed sensitivity analyses to assess the extent of depletion-of-susceptibles and the effect of excluding participants who enrolled in the extension for the Delta period, or that did not enrol for the Omicron period. All the sensitivity analyses provided results that were similar to those presented here, but the estimates were more uncertain (see Tables B.1 through C.6). All the analyses were conducted with the use of Stata software, version 15.1 (StataCorp). The results were independently replicated with the use of R software, version 4.1.1, survival package v.3.2-13 (R Foundation for Statistical Computing). Our annotated code is available at https://github.com/SIREN-study/SARS-CoV-2-Immunity.

## RESULTS

### Study population

We included a total of 19,614 individuals who were participating in regular SIREN testing between September 2021 and February 2022. The cohort were predominantly female (84%), of white ethnicity (89%), with a median age of 47 (IQR 16), 74% reported no comorbidities, and 69% were employed in clinical roles (Supplementary Table i).

We analysed protection over two different time periods: Delta (07 September 2021 to 30 November 2021; n= 18,429) and Omicron (13 December 2021 to 28 February 2022; n= 16,458) (Table 1). Third-dose vaccinations were introduced into our cohort on 16 September 2021 and coverage was 95% by the end of the analysis period (Supplementary Figure1). On 15 September 2021, 29% had a history of SARS-CoV- 2 infection, of which 1,901 (65%) were presumed infected in the Wild-Type period, 688 (24%) in the Alpha variant period and 8 (0.3%) in the Delta variant period (prior to the start of this analysis).

In the previously uninfected cohort, the PCR testing frequency before and after V3 was every 15.7 days and 11.2 days respectively in the Delta period and 11.0 days and 8.6 days in the Omicron period. In the previously infected cohort, in the Delta period it was every 14.2 and 11.7 days before and after V3; in the Omicron period it was 11.3 and 8.9 days.

### Primary outcome

There were 2467 primary infections and 881 reinfections during the Omicron period, 278 primary infections and 85 reinfections during the Delta period. Cumulative incidence rates were higher against Omicron (primary: 43/10,000 person-days, reinfections: 32/10,000 person-days) than Delta (primary: 4/10,000 person-days, reinfections: 1/10,000 person-days).

### Relative vaccine effectiveness following third dose in previously uninfected participants

In the previously uninfected cohort, third doses increased protection against both variants relative to waned dose-2 but were less effective against Omicron (Vaccine Effectiveness (VE) 0-2 months post-V3, 3-doses BNT162b2: 35% (95% Confidence Interval (CI) 21%-47%) vs. 63% (CI 40%-77%) than Delta (table 1). Heterologous third doses (BNT162b2, 2-dose ChAdOX1) were especially effective; with VE 0-2 months post-V3 over ≥68% for both variants. Dose-3 protection waned rapidly against Omicron, with no additional benefit >4-months.

### Protection from previous infection

When included as a marginal predictor, previous infection continued to provide additional protection against Omicron (67% (CI 56%-75%) 3-6 months post-infection), but this waned after 9 months to around 25%, markedly lower than against Delta where it remained at 85% (CI 81% - 89%) after more than 15 months (table 2). We found no evidence of interaction between vaccine-acquired and infection-acquired protection.

### Relative protection following infection and vaccination (hybrid protection)

In the previously infected cohort, with participants with two waned BNT162b2 doses and no previous infections as the reference group, we saw additional benefit associated with previous infection (table 3). Against Delta, a third dose and previous infection were associated with around 90% additional protection, even more than one year after primary infection. Against Delta, the combined additional protection remained around 70% for previous infection less than one year ago and around 50% for previous infection more than one year ago. Despite wide confidence intervals in the 4-month post-V3 category (reflecting small numbers of participants), there is an indication that the additional protective effect associated with previous infection wanes over time against Omicron, as in Table 2.

## DISCUSSION

In the wake of surging infections in the SIREN cohort driven by the Omicron-variant, deployment of third doses of BNT162b2 vaccines provided short-term protection against infection but this waned rapidly. Vaccinated and previously infected participants continued to have higher protection than those without previous infection, but this reduced with time since infection in the Omicron period. Protection in the Omicron period contrasted sharply with that of the Delta period, from both vaccines and previous infection, with both providing more modest impact against Omicron infections.

Short-term increased protection following vaccine third doses against Omicron infections, including symptomatic infections and severe illness, has been observed in other studies^5, 17–19^. Others have also reported reduced vaccine effectiveness against Omicron compared to Delta, and rapid waning.^7, 8, 18, 20, 21^ Our finding of more pronounced benefit following third doses among ChAdOX1 primary course recipients who received a heterologous third dose (BNT162b2), has been observed in both trials and real-world studies, related to the lower effectiveness of the ChAdOX1 primary series.^10, 18^

For previous strains of SARS-CoV-2, hybrid immunity offers greater and more durable protection against reinfection than infection or vaccination alone, with protection remaining >90% more than a year after primary infection.^6^ In this current analysis, this trend continued whilst the Delta-variant dominated, and then diverged with the emergence of Omicron. Whilst during the Omicron period the previously infected cohort maintained higher protection against (re)infection than the previously uninfected cohort, the impact was reduced, and showed evidence of waning just nine months after infection. Given the distinct SARS-CoV-2 variant waves experienced by our cohort,^22^ participants in the Omicron analysis who had been infected less than nine months previously were most likely infected by Delta-variant, those infected 9- 12 months ago had Alpha-variant infections, and those infected over 15 months ago had Wild-type infections. Possible explanations for this lower protection afforded by a primary infection sustained over 9 months ago could be greater antigenic distance between successive variants, with a less effective neutralisation response to Omicron from a wildtype infection than a Delta infection, or antibody waning over time.^12, 23–26^

The continued additional protection from hybrid immunity compared to vaccine alone suggests that there are alternative immunological pathways which confer greater protection. Most notable are differences with neutralising antibody (nAb) titres and breadth of neutralisation with hybrid immunity compared to vaccine or infection induced immunity.^27–33^ There are other immunological differences between the groups, including in tissue-resident memory T-cells, and the mucosal immune system,^33–36^ which has prompted interest in development of intranasal vaccination.^37^

The key strengths of our study include the size of our cohort undergoing frequent PCR testing independent of disease status, giving us confidence that most infections were detected. Having monitored our well-characterised cohort for infections and vaccination since June 2020, we have a valuable history of antigenic exposures for all our participants, enabling analysis of time since infection. The speed of third-dose vaccine deployment and frequent testing, independent of symptoms, considerably limited the effect of the depletion-of-susceptibles bias (supplementary appendix B). ^38^ This is especially relevant for Omicron, which is more often associated with less severe or asymptomatic disease,^39^ therefore infections are more likely to be missed, putting studies without frequent and asymptomatic testing at risk from the bias.^40^ In our cohort, frequency of testing during the Omicron period was high (every 8-9 days), therefore our estimates – and importantly the waning highlighted – are unlikely to be affected. Given the early start of third-dose deployment in our cohort, we were able to estimate the effectiveness of the third dose in the Delta dominant period in addition to the Omicron period, and the contrasting results powerfully demonstrate the step-change introduced by the Omicron-variant. The unspecified and varying baseline hazard rate of the Cox model is crucial to obtain reliable estimates in a period like winter 2021-2022, with a rapidly evolving wave of infections and changing recommendations for social distancing.

The most important limitation in our study is that we were unable to estimate absolute vaccine effectiveness and protection as too few participants remained unvaccinated to provide the reference group in this analysis. However, following the success of COVID-19 vaccine campaigns in high-income countries, this challenge is not unique to SIREN, and moreover, our selection of waned dose-2 participants as our reference group was appropriate given the decision to deploy third doses was in response to evidence of primary course vaccine waning. A further limitation is that testing tended to be less frequent before V3 than after; this might have led to underestimating V3 effectiveness.

Mass deployment of BNT162b2 vaccine third doses in response to the Omicron surge provided short-term protection against infection, and therefore helped mitigate record levels of SARS-CoV-2 infections in the UK. The rapid subsequent waning of vaccine protection is a significant challenge for future sustainable control measures, and future COVID-19 vaccine policy. Hybrid-immunity is increasingly commonplace. Our findings of reduced protection from hybrid-immunity in the Omicron era demonstrate the complexity of an evolving pandemic with potential emergence of immune-escape variants. This underscores the importance of continued longitudinal studies with well-characterised cohorts and reliable antigenic history.

### Data sharing

The metadata will be available through the Health Data Research UK Co-Connect platform. Anonymised data will be made available for secondary analysis to trusted researchers upon reasonable request.

### Statement of contributions

VH, FI and SF are joint first authors. VH, FI, SF, AC and SH designed the analysis plan. VH, FI and SF wrote the first draft. SF led on data management, cleaning and descriptive analysis. FI designed and undertook the statistical analysis, including sensitivity analyses, with statistical oversight provided by AC. PK independently replicated the statistical analysis in an alternative programme (R). SF, FI, PK and VH had full access to and analysed the data for this analysis. DS undertook and wrote the literature review. SF, AA, MC, JI, LP, DC, EL, CSB, are responsible for study delivery, oversight and ongoing data collection. SH is the Chief Investigator for the SIREN study and VH is the study lead, and guarantors of the study. All authors reviewed the draft manuscript and approved the final version for submission. All authors had full access to all the data in the study and accept responsibility to submit for publication.

## Supporting information

Strobe Checklist

---

**Figure 1a:**
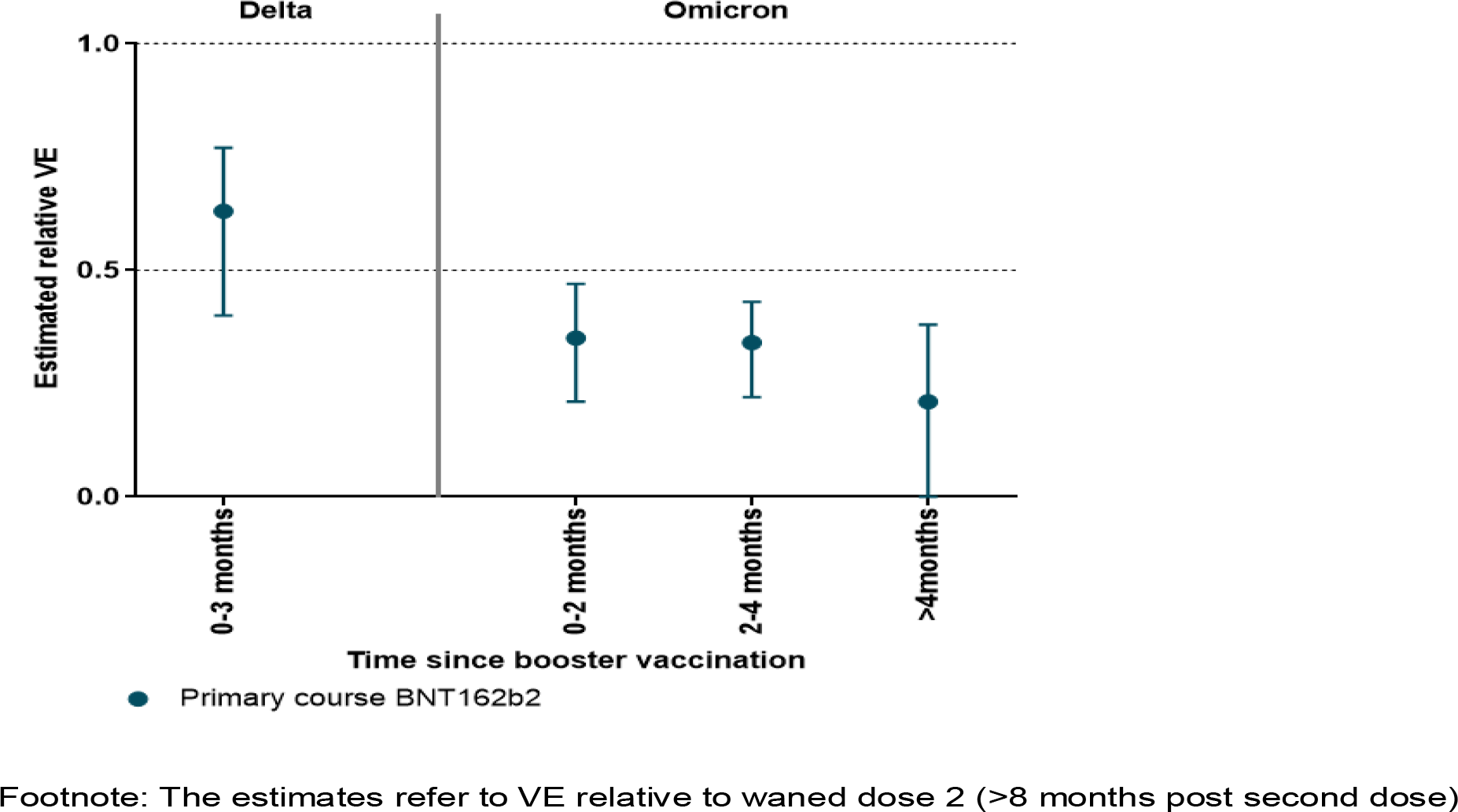
**Relative vaccine effectiveness following BNT162b2 V3 after BNT162b2 primary course by time since vaccination, against Delta-variant and Omicron-variant infections, among previously unifected cohort**

**Figure 1b:**
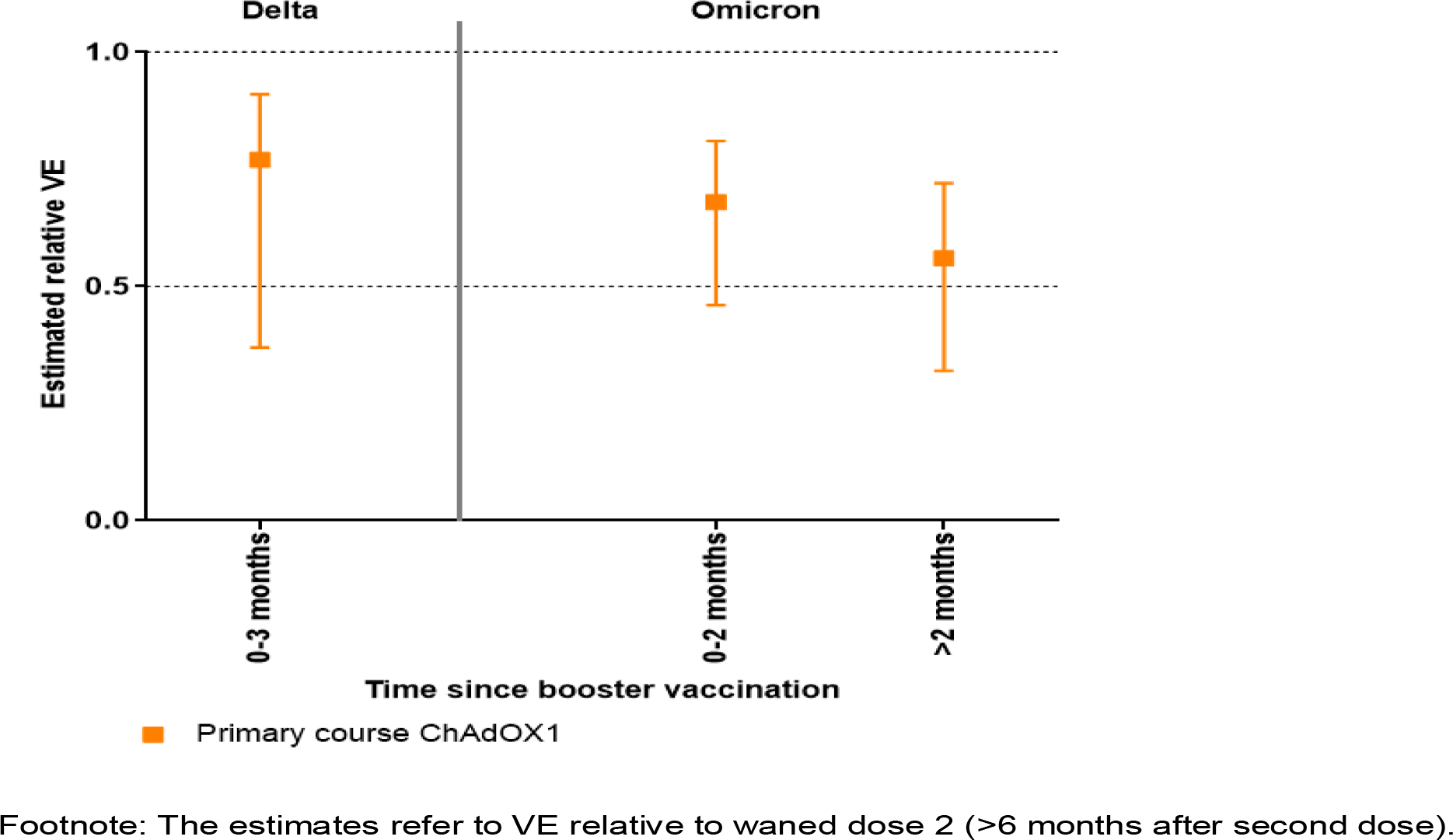
**Relative vaccine effectiveness following BNT162b2 V3 after ChAdOX1 primary course by time since vaccination, against Delta-variant and Omicron-variant infections, among previously unifected cohort**

**Figure 2:**
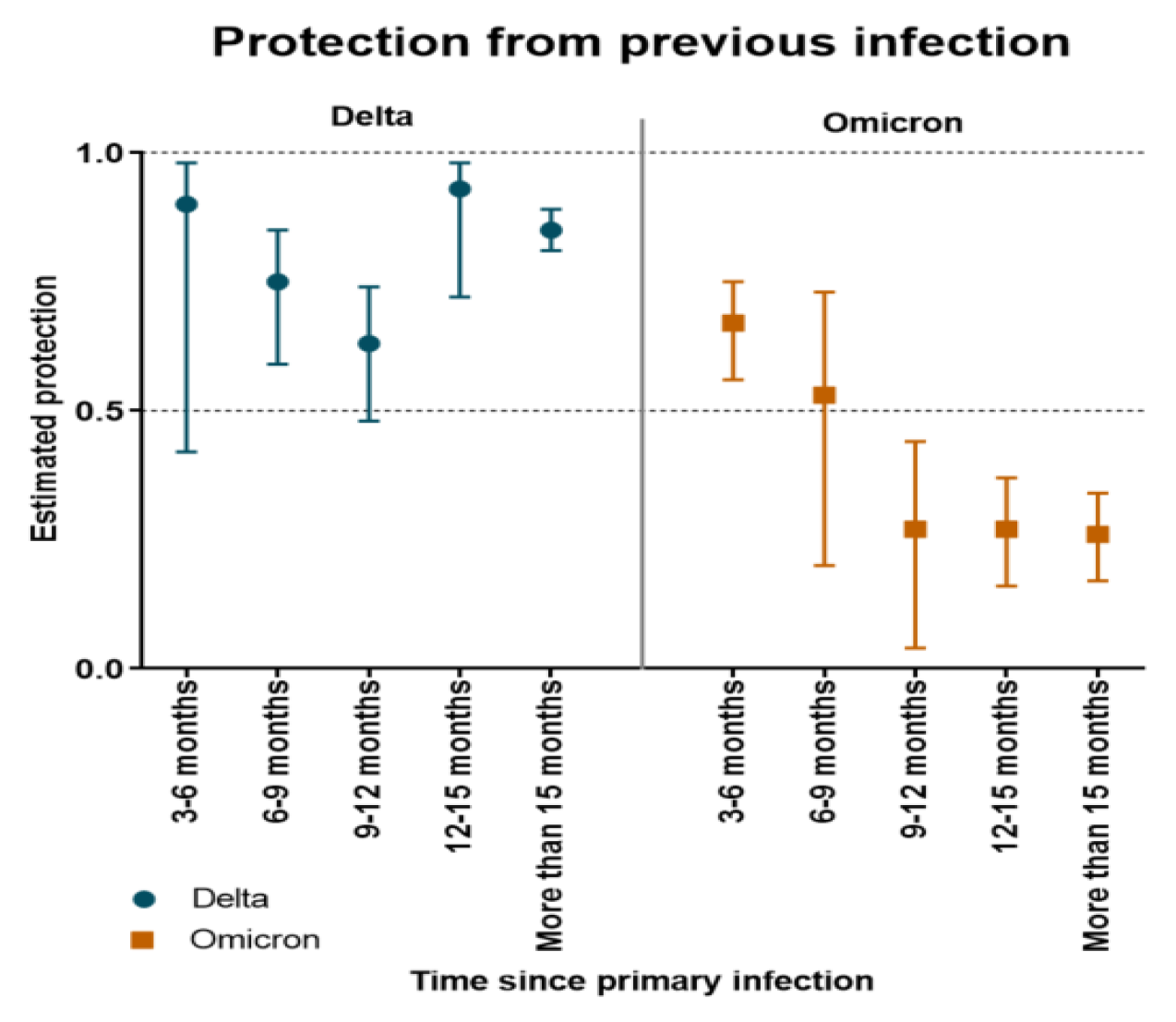
**Protection from infection over time, controlled for vaccine status, during Delta-variant and Omicron-variant periods**

**Figure 3:**
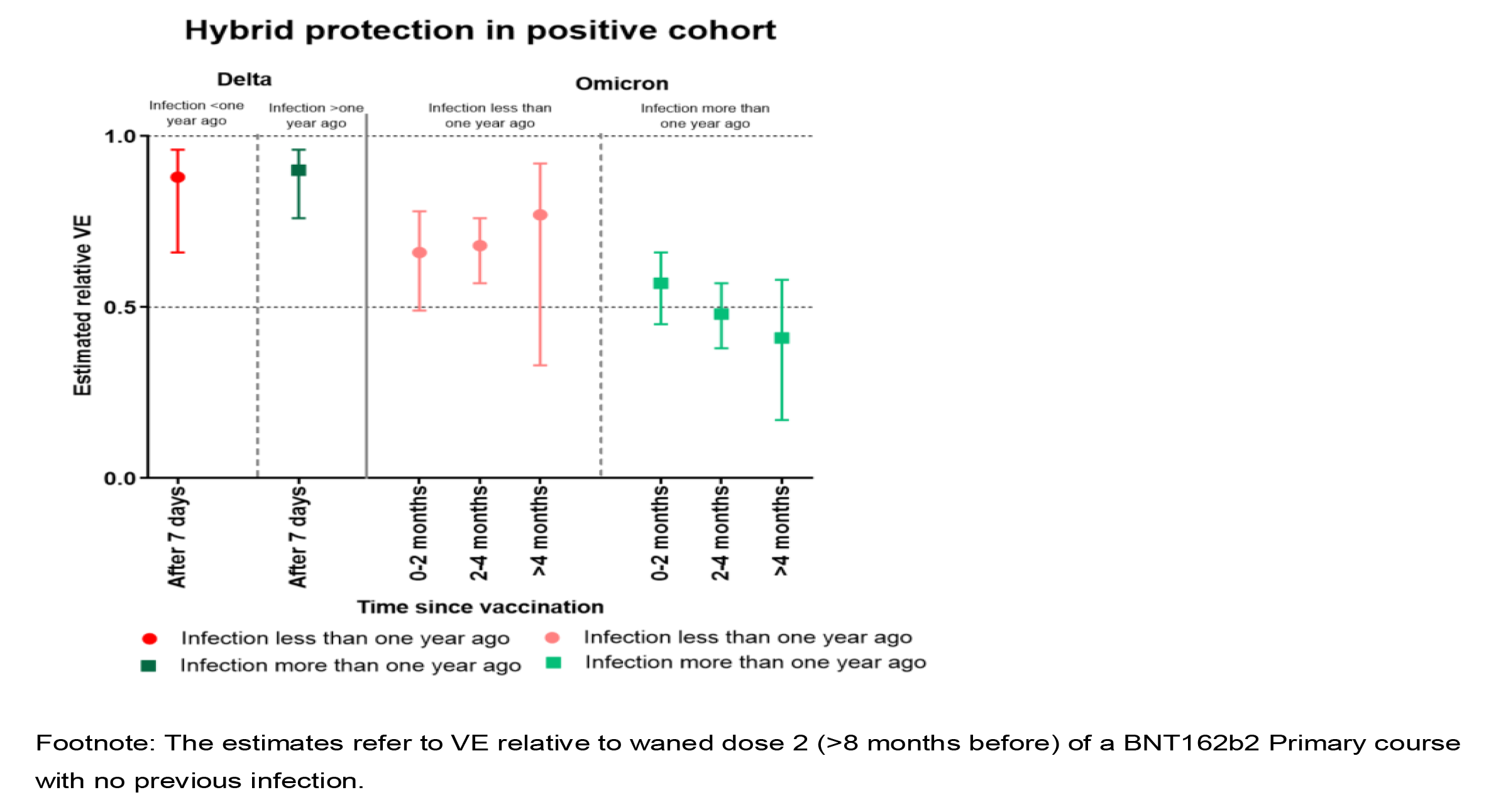
**Protection from infection and BNT162b2 third-vaccine dose (hybrid protection) during Delta-variant and Omicron-variant periods**

