## Supplementary Materials for "Effectiveness of BNT162b2 mRNA vaccine third doses and previous infection in protecting against SARS-CoV-2 infections during the Delta and Omicron variant waves; the UK SIREN cohort study September 2021 to February 2022"

### **Supplementary Table i: SIREN Study Group**

| **Organisation Name** | **First Name** | **Surname** |
| --- | --- | --- |
| UK Health Security Agency | Susan | Hopkins |
| UK Health Security Agency | Victoria | Hall |
| UK Health Security Agency | Jasmin | Islam |
| UK Health Security Agency | Ana | Atti |
| UK Health Security Agency | Omoyeni | Adebiyi |
| UK Health Security Agency | Nick | Andrews |
| UK Health Security Agency | Hannah | Emmett |
| UK Health Security Agency | Jonathan | Broad |
| UK Health Security Agency | Nish | Kapirial |
| UK Health Security Agency | Simone | Dyer |
| UK Health Security Agency | Sophie | Russell |
| UK Health Security Agency | Colin | Brown |
| UK Health Security Agency | Joanna | Conneely |
| UK Health Security Agency | Paul | Conneely |
| UK Health Security Agency | Sarah | Foulkes |
| UK Health Security Agency | Nabila | Fowles-Gutierrez |
| UK Health Security Agency | Nipunadi | Hettiarachchi |
| UK Health Security Agency | Jameel | Khawam |
| UK Health Security Agency | Edward | Monk |
| UK Health Security Agency | Katie | Munro |
| UK Health Security Agency | Andrew | Taylor-Kerr |
| UK Health Security Agency | Jean | Timeyin |
| UK Health Security Agency | Edgar | Wellington |
| UK Health Security Agency | Angela | Dunne |
| UK Health Security Agency | Dominic | Sparkes |
| UK Health Security Agency | Naomi | Platt |
| UK Health Security Agency | Anna | Howells |
| UK Health Security Agency | Enemona | Adaji |
| UK Health Security Agency | Omolola | Akinbami |
| UK Health Security Agency | Palak | Joshi |
| UK Health Security Agency | Paola | Barbero |
| UK Health Security Agency | Meera | Chand |
| UK Health Security Agency | Andre | Charlett |
| UK Health Security Agency | Michelle | Cole |
| UK Health Security Agency | Claire | Neill |
| UK Health Security Agency | Anne-Marie | O’Connell |
| UK Health Security Agency | Ferdinando | Insalata |
| UK Health Security Agency | Tim | Brooks |
| UK Health Security Agency | Maria | Zambon |
| UK Health Security Agency | Mary | Ramsay |
| UK Health Security Agency | Ayoub | Saei |
| UK Health Security Agency | Ezra | Linley |
| UK Health Security Agency | Simon | Tonge |
| UK Health Security Agency | Ashley | Otter |
| UK Health Security Agency | Silvia | D’Arcangelo |
| UK Health Security Agency | Cathy | Rowe |
| UK Health Security Agency | Amanda | Semper |
| UK Health Security Agency | Eileen | Gallagher |
| UK Health Security Agency | Robert | Kyffin |
| UK Health Security Agency | Kate | Howell |
| UK Health Security Agency | Jacqueline | Hewson |
| UK Health Security Agency | Iain | Milligan |
| UK Health Security Agency | Noshin | Sajedi |
| UK Health Security Agency | Davina | Calbraith |
| UK Health Security Agency | Caio | Tranquillini |
| UK Health Security Agency | Jerry | Ye Aung Kyaw |
| Public Health Agency Northern Ireland | Dianne | Corrigan |
| Public Health Agency Northern Ireland | Lisa | Cromey |
| Glasgow Caledonian University & Public Health Scotland | Lesley | Price |
| Glasgow Caledonian University & Public Health Scotland | Nicole | Sergenson |
| Glasgow Caledonian University & Public Health Scotland | Sally | Stewart |
| Glasgow Caledonian University & Public Health Scotland | Lynne | Haahr |
| Glasgow Caledonian University & Public Health Scotland | Desy | Nuryunarsih |
| Glasgow Caledonian University | Annelysse | Jorgenson |
| Glasgow Caledonian University | Ayodeji | Matuluko |
| Glasgow Caledonian University | Melanie | Dembinsky |
| Glasgow Caledonian University | Desmond | Areghan |
| Glasgow Caledonian University | Alexander | Olaoye |
| Public Health Scotland | Josie | Evans |
| Public Health Scotland | Jennifer | Bishop |
| Public Health Scotland | Jennifer | Weir |
| Public Health Scotland | Laura | Dobbie |
| Public Health Scotland | Andrew | Telfer |
| Public Health Scotland | David | Goldberg |
| University of St Andrews | David | Crossman |
| Public Health Scotland | Caitlin | Plank |
| Public Health Scotland | Laura | Naismith |
| Public Health Wales | Ellen | De Lacy |
| Public Health Wales | Guy | Stevens |
| Public Health Wales | Susannah | Froude |
| Public Health Wales | Linda | Tyson |
| Health and Care Research Wales | Yvette | Ellis |
| Health and Care Research Wales | Chris | Norman |
| **Participating NHS SIREN Sites** |  |  |
| ALDER HEY CHILDREN'S NHS FOUNDATION TRUST | Stephen | McWilliam |
| ALDER HEY CHILDREN'S NHS FOUNDATION TRUST | Beatriz | Larru |
| ASHFORD AND ST PETER'S HOSPITALS NHS FOUNDATION TRUST | Stephen | Winchester |
| ASHFORD AND ST PETER'S HOSPITALS NHS FOUNDATION TRUST | Samuel | Rowley |
| BASILDON AND THURROCK UNIVERSITY HOSPITALS NHS FOUNDATION TRUST | Stacey | Pepper |
| BASILDON AND THURROCK UNIVERSITY HOSPITALS NHS FOUNDATION TRUST | Georgina | Butt |
| BEDFORDSHIRE HOSPITALS NHS FOUNDATION TRUST | Simantee | Guha |
| BEDFORDSHIRE HOSPITALS NHS FOUNDATION TRUST | Philippa | Bakker |
| BELFAST HEALTH & SOCIAL CARE TRUST | Clodagh | Loughrey |
| BLACK COUNTRY HEALTHCARE NHS FOUNDATION TRUST | Alison | Grant |
| BLACK COUNTRY HEALTHCARE NHS FOUNDATION TRUST | Rebecca | Temple-Purcell |
| BLACKPOOL TEACHING HOSPITALS NHS FOUNDATION TRUST | Joanne | Howard |
| BLACKPOOL TEACHING HOSPITALS NHS FOUNDATION TRUST | Emma | Ward |
| BRIGHTON AND SUSSEX UNIVERSITY HOSPITALS NHS TRUST | Marion | Campbell |
| BUCKINGHAMSHIRE HEALTHCARE NHS TRUST | Nick | Wong |
| BUCKINGHAMSHIRE HEALTHCARE NHS TRUST | Ruth | Penn |
| CALDERDALE AND HUDDERSFIELD NHS FOUNDATION TRUST | N | Wong |
| CALDERDALE AND HUDDERSFIELD NHS FOUNDATION TRUST | G | Boyd |
| CHESTERFIELD ROYAL HOSPITAL NHS FOUNDATION TRUST | Edward | Harris |
| CHESTERFIELD ROYAL HOSPITAL NHS FOUNDATION TRUST | Amanda | Whileman |
| CORNWALL PARTNERSHIP NHS FOUNDATION TRUST | Susan | Greenwood |
| CORNWALL PARTNERSHIP NHS FOUNDATION TRUST | Angela | Pengilly |
| COUNTESS OF CHESTER HOSPITAL NHS FOUNDATION TRUST | Kim | Wells |
| COUNTESS OF CHESTER HOSPITAL NHS FOUNDATION TRUST | T | Barnes |
| CROYDON HEALTH SERVICES NHS TRUST | C | Jones |
| CROYDON HEALTH SERVICES NHS TRUST | Banerjee | Subhro-Osuji |
| CWM TAF MORGANNWG UNIVERSITY LHB | John | Geen |
| CWM TAF MORGANNWG UNIVERSITY LHB | Carla | Pothecary |
| DARTFORD AND GRAVESHAM NHS TRUST | Tracy | Edmunds |
| DARTFORD AND GRAVESHAM NHS TRUST | Nihil | Chitalia |
| DONCASTER AND BASSETLAW TEACHING HOSPITALS NHS FOUNDATION TRUST | Anna | Grice |
| DONCASTER AND BASSETLAW TEACHING HOSPITALS NHS FOUNDATION TRUST | K | Agwuh |
| DORSET COUNTY HOSPITAL NHS FOUNDATION TRUST | Jennifer | Graves |
| DORSET HEALTHCARE UNIVERSITY NHS FOUNDATION TRUST | James | Colton |
| DORSET HEALTHCARE UNIVERSITY NHS FOUNDATION TRUST | Stephanie | Willshaw |
| EAST SUSSEX HEALTHCARE NHS TRUST | Anna | Cowley |
| EAST SUSSEX HEALTHCARE NHS TRUST | Janet | Sinclair |
| EPSOM AND ST HELIER UNIVERSITY HOSPITALS NHS TRUST | Helen | Johnstone |
| EPSOM AND ST HELIER UNIVERSITY HOSPITALS NHS TRUST | Neringa | Vilimiene |
| FRIMLEY HEALTH NHS FOUNDATION TRUST | Manjula | Meda |
| FRIMLEY HEALTH NHS FOUNDATION TRUST | Jane | Democratis |
| GEORGE ELIOT HOSPITAL NHS TRUST | Simon | Brake |
| GEORGE ELIOT HOSPITAL NHS TRUST | David | Boss |
| GLOUCESTERSHIRE HOSPITALS NHS FOUNDATION TRUST | Amanda | Selassie |
| GREAT WESTERN HOSPITALS NHS FOUNDATION TRUST | Badrinathan | Chandrasekaran |
| GREAT WESTERN HOSPITALS NHS FOUNDATION TRUST | Eva | Fraile |
| HOUNSLOW AND RICHMOND COMMUNITY HEALTHCARE NHS TRUST | Shekoo | Mackay |
| HOUNSLOW AND RICHMOND COMMUNITY HEALTHCARE NHS TRUST | Shivani | Khan |
| HULL UNIVERSITY TEACHING HOSPITALS NHS TRUST | Phillipa | Burns |
| HULL UNIVERSITY TEACHING HOSPITALS NHS TRUST | Nicholas | Easom |
| HYWEL DDA UNIVERSITY LHB | Tracy | Lewis |
| IMPERIAL COLLEGE HEALTHCARE NHS TRUST | Graham | Pickard |
| IMPERIAL COLLEGE HEALTHCARE NHS TRUST | Keneisha | Lewis |
| JAMES PAGET UNIVERSITY HOSPITALS NHS FOUNDATION TRUST | Davis | Nwaka |
| JAMES PAGET UNIVERSITY HOSPITALS NHS FOUNDATION TRUST | Christian | Hacon |
| KING'S COLLEGE HOSPITAL NHS FOUNDATION TRUST | Ray | Chaudhuri |
| KING'S COLLEGE HOSPITAL NHS FOUNDATION TRUST | Jonnie | Aeron-Thomas |
| LEEDS TEACHING HOSPITALS NHS TRUST | Kyra | Holliday |
| LEEDS TEACHING HOSPITALS NHS TRUST | Clair | Favager |
| LEWISHAM AND GREENWICH NHS TRUST | A | Shah |
| LIVERPOOL UNIVERSITY HOSPITALS NHS FOUNDATION TRUST | Anu | Chawla |
| LIVERPOOL UNIVERSITY HOSPITALS NHS FOUNDATION TRUST | Fran | Westwell |
| LONDON NORTH WEST UNIVERSITY HEALTHCARE NHS TRUST | Ekaterina | Watson |
| MANCHESTER UNIVERSITY NHS FOUNDATION TRUST | Alexander | Horsley |
| MANCHESTER UNIVERSITY NHS FOUNDATION TRUST | Shazaad | Ahmad |
| MID ESSEX HOSPITAL SERVICES NHS TRUST | Lauren | Sach |
| MID ESSEX HOSPITAL SERVICES NHS TRUST | Yvonne | Lester |
| MID YORKSHIRE HOSPITALS NHS TRUST | Ismaelette | Del Rosario |
| MID YORKSHIRE HOSPITALS NHS TRUST | John | Ashcroft |
| MOORFIELDS EYE HOSPITAL NHS FOUNDATION TRUST | Roxanne | Crosby-Nwaobi |
| MOORFIELDS EYE HOSPITAL NHS FOUNDATION TRUST | Chloe | Reeks |
| NHS BORDERS | Joy | Dawson |
| NHS BORDERS | Lauren | Finlayson |
| NHS FIFE | Devesh | Dhasmana |
| NHS FIFE | Susan | Fowler |
| NHS FORTH VALLEY | Euan | Cameron |
| NHS FORTH VALLEY | Anne | Todd |
| NHS GREATER GLASGOW AND CLYDE | Antonia | Ho |
| NHS GREATER GLASGOW AND CLYDE | Michael | Murphy |
| NHS WESTERN ISLES | Martin | Malcolm |
| NHS WESTERN ISLES | Beth | Smith |
| NORFOLK AND NORWICH UNIVERSITY HOSPITALS NHS FOUNDATION TRUST | Ngozi | Elumogo |
| NORFOLK AND NORWICH UNIVERSITY HOSPITALS NHS FOUNDATION TRUST | Louise | Coke |
| NORTH CUMBRIA INTEGRATED CARE NHS FOUNDATION TRUST | Beverley | Wilkinson |
| NORTH CUMBRIA INTEGRATED CARE NHS FOUNDATION TRUST | John | Elliott |
| NORTH WEST ANGLIA NHS FOUNDATION TRUST | Janki | Bhayani |
| NORTH WEST ANGLIA NHS FOUNDATION TRUST | Stephanie | Diaz |
| NORTHERN DEVON HEALTHCARE NHS TRUST | M | Howard |
| NORTHERN HEALTH & SOCIAL CARE TRUST | Elinor | Hanna |
| NORTHERN HEALTH & SOCIAL CARE TRUST | Frances | Johnston |
| NORTHERN LINCOLNSHIRE AND GOOLE NHS FOUNDATION TRUST | Jonathan | Hatton |
| NORTHERN LINCOLNSHIRE AND GOOLE NHS FOUNDATION TRUST | Peter | Cowling |
| NOTTINGHAM UNIVERSITY HOSPITALS NHS TRUST | Sarah | Brand |
| NOTTINGHAM UNIVERSITY HOSPITALS NHS TRUST | Jack | Squires |
| PORTSMOUTH HOSPITALS NHS TRUST | Johanna | Mouland |
| PORTSMOUTH HOSPITALS NHS TRUST | Karen | Hudson |
| POWYS TEACHING LHB | Jayne | Goodwin |
| POWYS TEACHING LHB | Chris | Norman |
| QUEEN VICTORIA HOSPITAL NHS FOUNDATION TRUST | J | Giles |
| ROYAL BERKSHIRE NHS FOUNDATION TRUST | Maya | Joseph |
| ROYAL BERKSHIRE NHS FOUNDATION TRUST | Holly | Coles |
| ROYAL CORNWALL HOSPITALS NHS TRUST | H | Chenoweth |
| ROYAL DEVON AND EXETER NHS FOUNDATION TRUST | Cressida | Auckland |
| ROYAL DEVON AND EXETER NHS FOUNDATION TRUST | Stephanie | Prince |
| ROYAL NATIONAL ORTHOPAEDIC HOSPITAL NHS TRUST | Simon | Warren |
| ROYAL NATIONAL ORTHOPAEDIC HOSPITAL NHS TRUST | Esther | Hanison |
| ROYAL PAPWORTH HOSPITAL NHS FOUNDATION TRUST | Sumita | Pai |
| ROYAL PAPWORTH HOSPITAL NHS FOUNDATION TRUST | Allison | Doel |
| ROYAL SURREY COUNTY HOSPITAL NHS FOUNDATION TRUST | Charles | Piercy |
| ROYAL SURREY COUNTY HOSPITAL NHS FOUNDATION TRUST | Esther | Tarr |
| SHEFFIELD CHILDREN'S NHS FOUNDATION TRUST | James | Pethick |
| SHEFFIELD CHILDREN'S NHS FOUNDATION TRUST | S | Gormley |
| SHERWOOD FOREST HOSPITALS NHS FOUNDATION TRUST | Lynne | Allsop |
| SHERWOOD FOREST HOSPITALS NHS FOUNDATION TRUST | Shrikant | Ambalkar |
| SHROPSHIRE COMMUNITY HEALTH NHS TRUST | Johanne | Tomlinson |
| SOLENT NHS TRUST | Cathy | Price |
| SOMERSET NHS FOUNDATION TRUST | Justin | Pepperell |
| SOMERSET NHS FOUNDATION TRUST | Kate | James |
| SOUTHERN HEALTH & SOCIAL CARE TRUST | Angel | Boulos |
| SOUTHERN HEALTH & SOCIAL CARE TRUST | Fiona | Thompson |
| SOUTHPORT AND ORMSKIRK HOSPITAL NHS TRUST | Katherine | Gray |
| SOUTHPORT AND ORMSKIRK HOSPITAL NHS TRUST | Kerryanne | Brown |
| ST GEORGE'S UNIVERSITY HOSPITALS NHS FOUNDATION TRUST | Tim | Planche |
| ST GEORGE'S UNIVERSITY HOSPITALS NHS FOUNDATION TRUST | Angela | Houston |
| ST HELENS AND KNOWSLEY TEACHING HOSPITALS NHS TRUST | Rowan | Pritchard-Jones |
| ST HELENS AND KNOWSLEY TEACHING HOSPITALS NHS TRUST | Diane | Wycherley |
| STOCKPORT NHS FOUNDATION TRUST | Barzo | Faris |
| STOCKPORT NHS FOUNDATION TRUST | Liane | Marsh |
| SURREY AND SUSSEX HEALTHCARE NHS TRUST | Kofi | Nimako |
| SURREY AND SUSSEX HEALTHCARE NHS TRUST | Simon | Bax |
| THE CLATTERBRIDGE CANCER CENTRE NHS FOUNDATION TRUST | Sheena | Khanduri |
| THE CLATTERBRIDGE CANCER CENTRE NHS FOUNDATION TRUST | Nagesh | Kalakonda |
| THE DUDLEY GROUP NHS FOUNDATION TRUST | Helen | Ashby |
| THE HILLINGDON HOSPITALS NHS FOUNDATION TRUST | Natasha | Mahabir |
| THE NEWCASTLE UPON TYNE HOSPITALS NHS FOUNDATION TRUST | Brendan | Payne |
| THE NEWCASTLE UPON TYNE HOSPITALS NHS FOUNDATION TRUST | Jayne | Harwood |
| THE PRINCESS ALEXANDRA HOSPITAL NHS TRUST | Kathryn | Court |
| THE PRINCESS ALEXANDRA HOSPITAL NHS TRUST | Nikki | White |
| THE ROBERT JONES AND AGNES HUNT ORTHOPAEDIC HOSPITAL NHS FOUNDATION TRUST | Ruth | Longfellow |
| THE ROYAL BOURNEMOUTH AND CHRISTCHURCH HOSPITALS NHS FOUNDATION TRUST | Mihye | Lee |
| THE ROYAL WOLVERHAMPTON NHS TRUST | Marie | Green |
| THE ROYAL WOLVERHAMPTON NHS TRUST | Lauren | Hughes |
| TORBAY AND SOUTH DEVON NHS FOUNDATION TRUST | Matthew | Halkes |
| TORBAY AND SOUTH DEVON NHS FOUNDATION TRUST | Pauline | Mercer |
| UNITED LINCOLNSHIRE HOSPITALS NHS TRUST | Alun | Roebuck |
| UNIVERSITY HOSPITAL SOUTHAMPTON NHS FOUNDATION TRUST | Eleri | Wilson-Davies |
| UNIVERSITY HOSPITALS BRISTOL AND WESTON NHS FOUNDATION TRUST | Rajeka | Lazarus |
| UNIVERSITY HOSPITALS BRISTOL AND WESTON NHS FOUNDATION TRUST | Aaran | Sinclair |
| UNIVERSITY HOSPITALS COVENTRY AND WARWICKSHIRE NHS TRUST | N | Aldridge |
| UNIVERSITY HOSPITALS COVENTRY AND WARWICKSHIRE NHS TRUST | Lisa | Berry |
| UNIVERSITY HOSPITALS OF DERBY AND BURTON NHS FOUNDATION TRUST | L | Berry |
| UNIVERSITY HOSPITALS OF DERBY AND BURTON NHS FOUNDATION TRUST | F | Game |
| UNIVERSITY HOSPITALS OF LEICESTER NHS TRUST | Christopher | Holmes |
| UNIVERSITY HOSPITALS OF LEICESTER NHS TRUST | Martin | Wiselka |
| UNIVERSITY HOSPITALS OF NORTH MIDLANDS NHS TRUST | Christopher | Duff |
| UNIVERSITY HOSPITALS OF NORTH MIDLANDS NHS TRUST | Martin | Booth |
| VELINDRE NHS TRUST | Charlotte | Young |
| VELINDRE NHS TRUST | James | Powell |
| WALSALL HEALTHCARE NHS TRUST | Lisa | Richardson |
| WALSALL HEALTHCARE NHS TRUST | Aiden | Plant |
| WARRINGTON AND HALTON TEACHING HOSPITALS NHS FOUNDATION TRUST | Zaman | Qazzafi |
| WARRINGTON AND HALTON TEACHING HOSPITALS NHS FOUNDATION TRUST | Lisa | Ditchfield |
| WEST SUFFOLK NHS FOUNDATION TRUST | Veronica | Mendez Moro |
| WEST SUFFOLK NHS FOUNDATION TRUST | A | Moody |
| WESTERN HEALTH & SOCIAL CARE TRUST | Tracy | Donaghy |
| WESTERN HEALTH & SOCIAL CARE TRUST | Maurice | O'Kane |
| WESTERN SUSSEX HOSPITALS NHS FOUNDATION TRUST | R | Sierra |
| WHITTINGTON HEALTH NHS TRUST | Philippa | Kemsley |
| WHITTINGTON HEALTH NHS TRUST | Zehra'a | Al-Khafaji |
| WIRRAL UNIVERSITY TEACHING HOSPITAL NHS FOUNDATION TRUST | D | Harvey |
| WIRRAL UNIVERSITY TEACHING HOSPITAL NHS FOUNDATION TRUST | Y | Huang |
| WYE VALLEY NHS TRUST | L | Robinson |
| YEOVIL DISTRICT HOSPITAL NHS FOUNDATION TRUST | Sarah | Board |
| YEOVIL DISTRICT HOSPITAL NHS FOUNDATION TRUST | Andrew | Broadley |
| YORK TEACHING HOSPITAL NHS FOUNDATION TRUST | Claire | Brookes |
| YORK TEACHING HOSPITAL NHS FOUNDATION TRUST | Neil | Todd |
| ANEURIN BEVAN UNIVERSITY LHB | John | Northfield |
| ANEURIN BEVAN UNIVERSITY LHB | Sean | Cutler |
| BETSI CADWALADR UNIVERSITY LHB | Christian | Subbe |
| BETSI CADWALADR UNIVERSITY LHB | Caroline | Mulvaney Jones |
| BIRMINGHAM AND SOLIHULL MENTAL HEALTH NHS FOUNDATION TRUST | Manny | Bagary |
| BIRMINGHAM AND SOLIHULL MENTAL HEALTH NHS FOUNDATION TRUST | Di | Baines |
| BIRMINGHAM COMMUNITY HEALTHCARE NHS FOUNDATION TRUST | Rebecca | Chapman |
| BIRMINGHAM COMMUNITY HEALTHCARE NHS FOUNDATION TRUST | Lucy | Booth |
| BOLTON NHS FOUNDATION TRUST | Chinari | Subudhi |
| BOLTON NHS FOUNDATION TRUST | Scott | Latham |
| CENTRAL AND NORTH WEST LONDON NHS FOUNDATION TRUST | Abigail | Severn |
| CENTRAL AND NORTH WEST LONDON NHS FOUNDATION TRUST | Alejandro | Arenas-Pinto |
| DERBYSHIRE COMMUNITY HEALTH SERVICES NHS FOUNDATION TRUST | Sarah | Creer |
| DERBYSHIRE COMMUNITY HEALTH SERVICES NHS FOUNDATION TRUST | Eve | Etell Kirby |
| DERBYSHIRE HEALTHCARE NHS FOUNDATION TRUST | Joely | Morgan |
| DERBYSHIRE HEALTHCARE NHS FOUNDATION TRUST | Gemma | Harrison |
| DEVON PARTNERSHIP NHS TRUST | Nicola | Walker |
| DEVON PARTNERSHIP NHS TRUST | Anna | Grice |
| EAST SUFFOLK AND NORTH ESSEX NHS FOUNDATION TRUST | Luke | Bedford |
| EAST SUFFOLK AND NORTH ESSEX NHS FOUNDATION TRUST | Paul | Ridley |
| GOLDEN JUBILEE NATIONAL HOSPITAL | Val | Irvine |
| GOLDEN JUBILEE NATIONAL HOSPITAL | Elizabeth | Boyd |
| HAMPSHIRE HOSPITALS NHS FOUNDATION TRUST | Claire | Thomas |
| HAMPSHIRE HOSPITALS NHS FOUNDATION TRUST | Ina | Hoad |
| ISLE OF WIGHT NHS TRUST | Emily | Macnaughton |
| ISLE OF WIGHT NHS TRUST | Sarah | Knight |
| LANCASHIRE & SOUTH CUMBRIA NHS FOUNDATION TRUST | Robert | Shorten |
| LANCASHIRE & SOUTH CUMBRIA NHS FOUNDATION TRUST | Kathryn | Hollinshead |
| LANCASHIRE TEACHING HOSPITALS NHS FOUNDATION TRUST | Robert | Shorten |
| LANCASHIRE TEACHING HOSPITALS NHS FOUNDATION TRUST | Claire | Corless |
| LEICESTERSHIRE PARTNERSHIP NHS TRUST | Sarah | Baillon |
| LEICESTERSHIRE PARTNERSHIP NHS TRUST | Samantha | Hamer |
| LINCOLNSHIRE PARTNERSHIP NHS FOUNDATION TRUST | Vijayendra | Waykar |
| LINCOLNSHIRE PARTNERSHIP NHS FOUNDATION TRUST | Rebecca | Rutter |
| MAIDSTONE AND TUNBRIDGE WELLS NHS TRUST | Maureen | Williams |
| MAIDSTONE AND TUNBRIDGE WELLS NHS TRUST | Bethany | Jones |
| MID CHESHIRE HOSPITALS NHS FOUNDATION TRUST | Elijah | Matovu |
| MID CHESHIRE HOSPITALS NHS FOUNDATION TRUST | Claire | Gabriel |
| NHS GRAMPIAN | Harriet | Carroll |
| NHS GRAMPIAN | Alison | Thornton |
| NHS HIGHLAND | Andrew | Gibson |
| NHS HIGHLAND | Alexandra | Cochrane |
| NHS LANARKSHIRE | Manish | Patel |
| NHS LANARKSHIRE | Berni | Welsh |
| NHS LOTHIAN | Kate | Templeton |
| NHS LOTHIAN | Sam | Donaldson |
| NORTH MIDDLESEX UNIVERSITY HOSPITAL NHS TRUST | Mariyam | Mirfenderesky |
| NORTH MIDDLESEX UNIVERSITY HOSPITAL NHS TRUST | Yasmin | Lahrach |
| POOLE HOSPITAL NHS FOUNDATION TRUST | Charlotte | Barclay |
| POOLE HOSPITAL NHS FOUNDATION TRUST | Liz | Sheridan |
| ROYAL FREE LONDON NHS FOUNDATION TRUST | Alison | Rodger |
| ROYAL FREE LONDON NHS FOUNDATION TRUST | Tabitha | Mahungu |
| ROYAL UNITED HOSPITALS BATH NHS FOUNDATION TRUST | Sarah | Meisner |
| ROYAL UNITED HOSPITALS BATH NHS FOUNDATION TRUST | Julia | Vasant |
| SALISBURY NHS FOUNDATION TRUST | Abby | Rand |
| SALISBURY NHS FOUNDATION TRUST | Catherine | Thompson |
| SANDWELL AND WEST BIRMINGHAM HOSPITALS NHS TRUST | Ash | Turner |
| SANDWELL AND WEST BIRMINGHAM HOSPITALS NHS TRUST | Anne | Hayes |
| SHEFFIELD TEACHING HOSPITALS NHS FOUNDATION TRUST | Thushan | de Silva |
| SHEFFIELD TEACHING HOSPITALS NHS FOUNDATION TRUST | Helen | Shulver |
| SHREWSBURY AND TELFORD HOSPITAL NHS TRUST | Mandy | Carnahan |
| SHREWSBURY AND TELFORD HOSPITAL NHS TRUST | Mandy | Beekes |
| SOUTH EASTERN HEALTH & SOCIAL CARE | Yuri | Protaschik |
| SOUTH EASTERN HEALTH & SOCIAL CARE | Susan | Regan |
| SOUTHEND UNIVERSITY HOSPITAL NHS FOUNDATION TRUST | John | Day |
| SOUTHEND UNIVERSITY HOSPITAL NHS FOUNDATION TRUST | Swapna | Kunhunny |
| SOUTHERN HEALTH NHS FOUNDATION TRUST | Alice | Neave |
| SOUTHERN HEALTH NHS FOUNDATION TRUST | James | Stanley-Watson |
| SWANSEA BAY UNIVERSITY LHB | Claire | Stafford |
| SWANSEA BAY UNIVERSITY LHB | Rebeccah | Thomas |
| UNIVERSITY HOSPITALS OF MORECAMBE BAY NHS FOUNDATION TRUST | Lynda | Fothergill |
| UNIVERSITY HOSPITALS OF MORECAMBE BAY NHS FOUNDATION TRUST | Karen | Burns |
| UNIVERSITY HOSPITALS PLYMOUTH NHS TRUST | David | Hilton |
| UNIVERSITY HOSPITALS PLYMOUTH NHS TRUST | Hannah | Jory |
| **SIREN Associated Studies** | **First name** | **Surname** |
| Protective Immunity from T cells to Covid-19 in Health workers (PITCH) | Susanna | Dunachie |
| Protective Immunity from T cells to Covid-19 in Health workers (PITCH) | Paul | Klenerman |
| Protective Immunity from T cells to Covid-19 in Health workers (PITCH) | Chris | Duncan |
| Protective Immunity from T cells to Covid-19 in Health workers (PITCH) | Rebecca | Payne |
| Protective Immunity from T cells to Covid-19 in Health workers (PITCH) | Lance | Turtle |
| Protective Immunity from T cells to Covid-19 in Health workers (PITCH) | Alex | Richter |
| Protective Immunity from T cells to Covid-19 in Health workers (PITCH) | Thushan | De Silva |
| Protective Immunity from T cells to Covid-19 in Health workers (PITCH) | Eleanor | Barnes |
| Protective Immunity from T cells to Covid-19 in Health workers (PITCH) | Daniel | Wootton |
| Vaccine Immunity, Breakthrough & Reinfection - Antibodies & T-cells (VIBRANT) Study | Oliver | Galgut |
| The Humoral Immune Correlates for COVID-19 (HICC) consortium | Jonathan | Heeney |
| The Humoral Immune Correlates for COVID-19 (HICC) consortium | Helen | Baxendale |
| The Humoral Immune Correlates for COVID-19 (HICC) consortium | Javier | Castillo-Olivares |
| The Francis Crick Institute | Rupert | Beale |
| The Francis Crick Institute | Edward | Carr |
| Genotype2Phenotype (G2P) Imperial College London | Wendy | Barclay |
| Genotype2Phenotype (G2P) Imperial College London | Maya | Moshe |
| Genotype2Phenotype (G2P) Glagow University | Massimo | Palmarini |
| Genotype2Phenotype (G2P) Glagow University | Brian | Willett |
| GenOMICC | John Kenneth | Baillie |

### **Supplementary Table ii: Demographics of participants by previous infection status (cohort), stratified by analysis period: Delta-variant dominant September 2021-November 2021, Omicron-dominant December 2021-February 2022**

|  | **Delta analysis** | | | | **Omicron analysis** | | | | ***P-value Delta vs Omicron*** | **Total all participants** |
| --- | --- | --- | --- | --- | --- | --- | --- | --- | --- | --- |
|  | **Previously uninfected cohort** | **Previously infected cohort** | ***P-value*** | **Total** | **Previously uninfected cohort** | **Previously infected cohort** | ***P-value*** | **Total** |  |  |
| **Gender** |  |  |  |  |  |  |  |  |  |  |
| Male | 2038 (15.7) | 981 (18.1) | *0.094* | **3019 (16.4)** | 1716 (15.5) | 946 (17.6) | *0.165* | **2662 (16.2)** | *0.833* | **3176 (16.2)** |
| Female | 10952 (84.2) | 4441 (81.9) | *<0.001* | **15393 (83.5)** | 9345 (84.4) | 4436 (82.4) | *0.003* | **13781 (83.7)** | *0.835* | **16420 (83.7)** |
| **Age group** |  |  |  |  |  |  |  |  |  |  |
| Under 25 | 308 (2.4) | 138 (2.5) | *0.911* | **446 (2.4)** | 270 (2.4) | 114 (2.1) | *0.849* | **384 (2.3)** | *0.024* | **496 (2.5)** |
| 25 to 34 | 2007 (15.4) | 897 (16.5) | *0.452* | **2904 (15.8)** | 1697 (15.3) | 869 (16.1) | *0.590* | **2566 (15.6)** | *0.158* | **3146 (16)** |
| 35 to 44 | 3157 (24.3) | 1327 (24.5) | *0.896* | **4484 (24.3)** | 2696 (24.3) | 1370 (25.4) | *0.442* | **4066 (24.7)** | *0.243* | **4875 (24.9)** |
| 45 to 54 | 4328 (33.3) | 1792 (33) | *0.850* | **6120 (33.2)** | 3673 (33.2) | 1810 (33.6) | *0.739* | **5483 (33.3)** | *0.332* | **6460 (32.9)** |
| 55 to 64 | 2953 (22.7) | 1199 (22.1) | *0.671* | **4152 (22.5)** | 2538 (22.9) | 1156 (21.5) | *0.328* | **3694 (22.4)** | *0.225* | **4308 (22)** |
| Over 65 | 251 (1.9) | 72 (1.3) | *0.734* | **323 (1.8)** | 200 (1.8) | 65 (1.2) | *0.743* | **265 (1.6)** | *0.018* | **329 (1.7)** |
| **Ethnicity** |  |  |  |  |  |  |  |  |  |  |
| White | 11707 (90) | 4692 (86.5) | *<0.001* | **16399 (89)** | 9960 (89.9) | 4661 (86.6) | *<0.001* | **14621 (88.8)** | *0.890* | **17401 (88.7)** |
| Asian | 800 (6.2) | 486 (9) | *0.059* | **1286 (7)** | 660 (6) | 463 (8.6) | *0.089* | **1123 (6.8)** | *0.070* | **1377 (7)** |
| Black | 161 (1.2) | 93 (1.7) | *0.757* | **254 (1.4)** | 154 (1.4) | 113 (2.1) | *0.657* | **267 (1.6)** | *0.014* | **308 (1.6)** |
| Mixed race | 169 (1.3) | 74 (1.4) | *0.968* | **243 (1.3)** | 151 (1.4) | 73 (1.4) | *0.996* | **224 (1.4)** | *0.013* | **263 (1.3)** |
| Other ethnic group | 148 (1.1) | 76 (1.4) | *0.866* | **224 (1.2)** | 131 (1.2) | 71 (1.3) | *0.933* | **202 (1.2)** | *0.012* | **240 (1.2)** |
| Prefer not to say | 19 (0.1) | 4 (0.1) | *0.971* | **23 (0.1)** | 18 (0.2) | 3 (0.1) | *0.964* | **21 (0.1)** | *0.001* | **25 (0.1)** |
| **Medical conditions category** |  |  |  |  |  |  |  |  |  |  |
| No medical condition | 9574 (73.6) | 4002 (73.8) | *0.860* | **13576 (73.7)** | 8207 (74.1) | 4020 (74.7) | *0.509* | **12227 (74.3)** | *0.737* | **14523 (74)** |
| Immunosuppression | 336 (2.6) | 101 (1.9) | *0.679* | **437 (2.4)** | 287 (2.6) | 98 (1.8) | *0.666* | **385 (2.3)** | *0.024* | **463 (2.4)** |
| Chronic respiratory conditions | 1631 (12.5) | 678 (12.5) | *0.976* | **2309 (12.5)** | 1368 (12.4) | 652 (12.1) | *0.876* | **2020 (12.3)** | *0.125* | **2425 (12.4)** |
| Chronic non-respiratory conditions | 1463 (11.3) | 644 (11.9) | *0.680* | **2107 (11.4)** | 1212 (10.9) | 614 (11.4) | *0.768* | **1826 (11.1)** | *0.114* | **2203 (11.2)** |
| **Staff group** |  |  |  |  |  |  |  |  |  |  |
| Administrative/Executive (office based) | 2202 (16.9) | 719 (13.3) | *0.020* | **2921 (15.9)** | 1908 (17.2) | 740 (13.7) | *0.029* | **2648 (16.1)** | *0.159* | **3128 (15.9)** |
| Doctor | 1674 (12.9) | 732 (13.5) | *0.678* | **2406 (13.1)** | 1387 (12.5) | 691 (12.8) | *0.841* | **2078 (12.6)** | *0.131* | **2491 (12.7)** |
| Nursing | 4207 (32.4) | 1936 (35.7) | *0.010* | **6143 (33.3)** | 3563 (32.2) | 1904 (35.4) | *0.017* | **5467 (33.2)** | *0.333* | **6548 (33.4)** |
| Healthcare Assistant | 827 (6.4) | 423 (7.8) | *0.341* | **1250 (6.8)** | 680 (6.1) | 406 (7.5) | *0.371* | **1086 (6.6)** | *0.068* | **1346 (6.9)** |
| Midwife | 251 (1.9) | 101 (1.9) | *0.966* | **352 (1.9)** | 202 (1.8) | 104 (1.9) | *0.947* | **306 (1.9)** | *0.019* | **374 (1.9)** |
| Healthcare Scientist | 612 (4.7) | 162 (3) | *0.340* | **774 (4.2)** | 533 (4.8) | 175 (3.3) | *0.383* | **708 (4.3)** | *0.042* | **829 (4.2)** |
| Pharmacist | 398 (3.1) | 126 (2.3) | *0.666* | **524 (2.8)** | 327 (3) | 123 (2.3) | *0.700* | **450 (2.7)** | *0.028* | **554 (2.8)** |
| Physiotherapist/Occupational Therapist/SALT | 571 (4.4) | 309 (5.7) | *0.390* | **880 (4.8)** | 479 (4.3) | 286 (5.3) | *0.533* | **765 (4.6)** | *0.048* | **937 (4.8)** |
| Student (Medical/Nursing/Midwifery/Other) | 231 (1.8) | 122 (2.2) | *0.760* | **353 (1.9)** | 225 (2) | 133 (2.5) | *0.784* | **358 (2.2)** | *0.019* | **381 (1.9)** |
| Estates/Porters/Security | 226 (1.7) | 114 (2.1) | *0.815* | **340 (1.8)** | 198 (1.8) | 115 (2.1) | *0.829* | **313 (1.9)** | *0.018* | **364 (1.9)** |
| Other | 1567 (12.1) | 622 (11.5) | *0.703* | **2189 (11.9)** | 1361 (12.3) | 636 (11.8) | *0.761* | **1997 (12.1)** | *0.119* | **2337 (11.9)** |
| Other without patient contact | 238 (1.8) | 59 (1.1) | *0.691* | **297 (1.6)** | 211 (1.9) | 71 (1.3) | *0.745* | **282 (1.7)** | *0.016* | **325 (1.7)** |
| **Occupational setting** |  |  |  |  |  |  |  |  |  |  |
| Office based | 2768 (21.3) | 922 (17) | *0.005* | **3690 (20)** | 2416 (21.8) | 979 (18.2) | *0.018* | **3395 (20.6)** | *0.200* | **3952 (20.1)** |
| Patient facing (non-clinical) | 566 (4.4) | 209 (3.9) | *0.759* | **775 (4.2)** | 494 (4.5) | 210 (3.9) | *0.737* | **704 (4.3)** | *0.042* | **833 (4.2)** |
| Outpatient | 3220 (24.8) | 1106 (20.4) | *0.003* | **4326 (23.5)** | 2700 (24.4) | 1106 (20.5) | *0.011* | **3806 (23.1)** | *0.235* | **4586 (23.4)** |
| Maternity/Labour Ward | 207 (1.6) | 73 (1.3) | *0.883* | **280 (1.5)** | 161 (1.5) | 69 (1.3) | *0.919* | **230 (1.4)** | *0.015* | **294 (1.5)** |
| Ambulance/Emergency Department/Inpatient Wards | 2385 (18.3) | 1442 (26.6) | *<0.001* | **3827 (20.8)** | 1916 (17.3) | 1286 (23.9) | *<0.001* | **3202 (19.5)** | *0.208* | **4058 (20.7)** |
| Intensive Care | 574 (4.4) | 194 (3.6) | *0.615* | **768 (4.2)** | 485 (4.4) | 192 (3.6) | *0.632* | **677 (4.1)** | *0.042* | **816 (4.2)** |
| Theatres | 452 (3.5) | 156 (2.9) | *0.718* | **608 (3.3)** | 367 (3.3) | 155 (2.9) | *0.796* | **522 (3.2)** | *0.033* | **639 (3.3)** |
| Other | 2832 (21.8) | 1323 (24.4) | *0.061* | **4155 (22.5)** | 2535 (22.9) | 1387 (25.8) | *0.044* | **3922 (23.8)** | *0.226* | **4436 (22.6)** |
| **Patient contact** |  |  |  |  |  |  |  |  |  |  |
| No | 10825 (83.2) | 4803 (88.5) | *<0.001* | **15628 (84.8)** | 9187 (83) | 4737 (88) | *<0.001* | **13924 (84.6)** | *0.848* | **16613 (84.7)** |
| Yes | 2179 (16.8) | 622 (11.5) | *0.001* | **2801 (15.2)** | 1887 (17) | 647 (12) | *0.003* | **2534 (15.4)** | *0.152* | **3001 (15.3)** |
| **Index of Multiple Deprivation** |  |  |  |  |  |  |  |  |  |  |
| 5 (least deprived) | 3509 (27) | 1359 (25.1) | *0.170* | **4868 (26.4)** | 2921 (26.4) | 1310 (24.3) | *0.159* | **4231 (25.7)** | *0.264* | **5101 (26)** |
| 4 | 2915 (22.4) | 1214 (22.4) | *0.979* | **4129 (22.4)** | 2479 (22.4) | 1154 (21.4) | *0.520* | **3633 (22.1)** | *0.224* | **4380 (22.3)** |
| 3 | 2564 (19.7) | 1093 (20.1) | *0.765* | **3657 (19.8)** | 2181 (19.7) | 1100 (20.4) | *0.618* | **3281 (19.9)** | *0.198* | **3903 (19.9)** |
| 2 | 2114 (16.3) | 885 (16.3) | *0.969* | **2999 (16.3)** | 1812 (16.4) | 907 (16.8) | *0.749* | **2719 (16.5)** | *0.163* | **3208 (16.4)** |
| 1 (most deprived) | 1189 (9.1) | 637 (11.7) | *0.078* | **1826 (9.9)** | 1010 (9.1) | 656 (12.2) | *0.045* | **1666 (10.1)** | *0.099* | **1999 (10.2)** |
| Not known | 713 (5.5) | 237 (4.4) | *0.503* | **950 (5.2)** | 671 (6.1) | 257 (4.8) | *0.450* | **928 (5.6)** | *0.052* | **1023 (5.2)** |
| **Region** |  |  |  |  |  |  |  |  |  |  |
| East Midlands | 835 (6.4) | 347 (6.4) | *0.987* | **1182 (6.4)** | 603 (5.4) | 289 (5.4) | *0.962* | **892 (5.4)** | *0.064* | **1250 (6.4)** |
| East of England | 1258 (9.7) | 627 (11.6) | *0.205* | **1885 (10.2)** | 1141 (10.3) | 634 (11.8) | *0.339* | **1775 (10.8)** | *0.102* | **1985 (10.1)** |
| London | 1300 (10) | 850 (15.7) | *<0.001* | **2150 (11.7)** | 1211 (10.9) | 902 (16.8) | *<0.001* | **2113 (12.8)** | *0.117* | **2330 (11.9)** |
| North East | 292 (2.2) | 184 (3.4) | *0.452* | **476 (2.6)** | 259 (2.3) | 182 (3.4) | *0.512* | **441 (2.7)** | *0.026* | **493 (2.5)** |
| North West | 1239 (9.5) | 721 (13.3) | *0.010* | **1960 (10.6)** | 1120 (10.1) | 736 (13.7) | *0.019* | **1856 (11.3)** | *0.106* | **2104 (10.7)** |
| South East | 948 (7.3) | 351 (6.5) | *0.609* | **1299 (7)** | 718 (6.5) | 309 (5.7) | *0.652* | **1027 (6.2)** | *0.071* | **1366 (7)** |
| South West | 1227 (9.4) | 310 (5.7) | *0.038* | **1537 (8.3)** | 939 (8.5) | 298 (5.5) | *0.098* | **1237 (7.5)** | *0.083* | **1622 (8.3)** |
| West Midlands | 767 (5.9) | 401 (7.4) | *0.322* | **1168 (6.3)** | 572 (5.2) | 367 (6.8) | *0.291* | **939 (5.7)** | *0.063* | **1264 (6.4)** |
| Yorkshire and Humber | 886 (6.8) | 444 (8.2) | *0.364* | **1330 (7.2)** | 714 (6.4) | 409 (7.6) | *0.464* | **1123 (6.8)** | *0.072* | **1413 (7.2)** |
| Scotland | 3154 (24.3) | 775 (14.3) | *<0.001* | **3929 (21.3)** | 2804 (25.3) | 816 (15.2) | *<0.001* | **3620 (22)** | *0.213* | **4181 (21.3)** |
| Northern Ireland | 645 (5) | 214 (3.9) | *0.543* | **859 (4.7)** | 619 (5.6) | 244 (4.5) | *0.532* | **863 (5.2)** | *0.047* | **928 (4.7)** |
| Wales | 453 (3.5) | 201 (3.7) | *0.888* | **654 (3.5)** | 374 (3.4) | 198 (3.7) | *0.852* | **572 (3.5)** | *0.036* | **678 (3.5)** |
| **Total** | **13004** | **5425** |  | **18429** | **11074** | **5384** |  | **16458** |  | **19614** |

**Note:** Participants recorded as non-binary or preferred not to disclose their gender have been removed from the table due to small number of participants (n=18)

### **Supplementary Figure 1: Proportion of vaccinated participants and variant time period, November 2020 to February 2022**

Proportion vaccinated

| 14 day period | Vaccine dose three | Vaccine dose two | Vaccine dose one | Alpha | Delta | Omicron BA.1 | Omicron BA.2 | Delta analysis period | Omicron analysis period |
| --- | --- | --- | --- | --- | --- | --- | --- | --- | --- |
| 2020-11-30 |  |  | 6.29512 | 105 |  |  |  |  |  |
| 2020-12-14 |  |  | 21.8937 | 105 |  |  |  |  |  |
| 2020-12-28 |  | 6.83701 | 51.2526 | 105 |  |  |  |  |  |
| 2021-01-11 |  | 8.87656 | 81.5904 | 105 |  |  |  |  |  |
| 2021-01-25 |  | 11.0259 | 84.793 | 105 |  |  |  |  |  |
| 2021-02-08 |  | 15.2976 | 81.8973 | 105 |  |  |  |  |  |
| 2021-02-22 |  | 31.7747 | 66.4369 | 105 |  |  |  |  |  |
| 2021-03-08 |  | 57.3272 | 41.7252 | 105 |  |  |  |  |  |
| 2021-03-22 |  | 84.419 | 15.0756 | 105 |  |  |  |  |  |
| 2021-04-05 |  | 94.4062 | 5.20084 | 105 |  |  |  |  |  |
| 2021-04-19 |  | 96.6656 | 3.00806 |  | 105 |  |  |  |  |
| 2021-05-03 |  | 97.7414 | 1.99857 |  | 105 |  |  |  |  |
| 2021-05-17 |  | 98.7611 | 1.06047 |  | 105 |  |  |  |  |
| 2021-05-31 |  | 99.3474 | 0.545529 |  | 105 |  |  |  |  |
| 2021-06-14 |  | 99.5615 | 0.387478 |  | 105 |  |  |  |  |
| 2021-06-28 |  | 99.6839 | 0.280412 |  | 105 |  |  |  |  |
| 2021-07-12 |  | 99.791 | 0.183542 |  | 105 |  |  |  |  |
| 2021-07-26 |  | 99.8521 | 0.132558 |  | 105 |  |  |  |  |
| 2021-08-09 |  | 99.8929 | 0.0968696 |  | 105 |  |  |  |  |
| 2021-08-23 |  | 99.9235 | 0.076476 |  | 105 |  |  |  |  |
| 2021-09-06 | 0.754563 | 99.2046 | 0.0407872 |  | 105 |  |  | 120 |  |
| 2021-09-20 | 34.9704 | 65.0041 | 0.0254998 |  | 105 |  |  | 120 |  |
| 2021-10-04 | 65.8923 | 34.0923 | 0.0153846 |  | 105 |  |  | 120 |  |
| 2021-10-18 | 78.7653 | 21.2347 | 0 |  | 105 |  |  | 120 |  |
| 2021-11-01 | 84.8238 | 15.1762 | 0 |  | 105 |  |  | 120 |  |
| 2021-11-15 | 87.7512 | 12.2488 | 0 |  | 105 |  |  | 120 |  |
| 2021-11-29 | 90.2218 | 9.77824 | 0 |  | 105 |  |  | 120 |  |
| 2021-12-13 | 92.8968 | 7.10321 | 0 |  |  | 105 |  |  | 120 |
| 2021-12-27 | 93.8955 | 6.10449 | 0 |  |  | 105 |  |  | 120 |
| 2022-01-10 | 94.4584 | 5.54159 | 0 |  |  | 105 |  |  | 120 |
| 2022-01-24 | 94.7985 | 5.20147 | 0 |  |  | 105 |  |  | 120 |
| 2022-02-07 | 94.9239 | 5.07605 | 0 |  |  |  | 105 |  | 120 |
| 2022-02-21 | 95.0982 | 4.90177 | 0 |  |  |  | 105 |  | 120 |

**Note:** Delta analysis period 07 September 2021 to 30 November 2021; Omicron analysis period 13 December 2021 to 28 February 2022.

### **Supplementary Figure 2: Number of PCR positive samples (primary infections and reinfection) in the previously uninfected and previously infected cohorts**

Nunber of infections

| Week | Primary infections | Reinfections | Delta | Omicron |
| --- | --- | --- | --- | --- |
| 2021-09-06 | 51 |  | 600 |  |
| 2021-09-13 | 31 |  | 600 |  |
| 2021-09-20 | 47 |  | 600 |  |
| 2021-09-27 | 50 | 1 | 600 |  |
| 2021-10-04 | 41 | 2 | 600 |  |
| 2021-10-11 | 33 | 3 | 600 |  |
| 2021-10-18 | 32 | 2 | 600 |  |
| 2021-10-25 | 18 | 0 | 600 |  |
| 2021-11-01 | 25 | 0 | 600 |  |
| 2021-11-08 | 17 | 0 | 600 |  |
| 2021-11-15 | 13 | 3 | 600 |  |
| 2021-11-22 | 4 | 4 | 600 |  |
| 2021-11-29 | 5 | 8 |  |  |
| 2021-12-06 | 2 | 10 |  |  |
| 2021-12-13 | 20 | 42 |  | 600 |
| 2021-12-20 | 42 | 99 |  | 600 |
| 2021-12-27 | 56 | 189 |  | 600 |
| 2022-01-03 | 53 | 146 |  | 600 |
| 2022-01-10 | 23 | 82 |  | 600 |
| 2022-01-17 | 28 | 63 |  | 600 |
| 2022-01-24 | 27 | 77 |  | 600 |
| 2022-01-31 | 19 | 68 |  | 600 |
| 2022-02-07 | 11 | 47 |  | 600 |
| 2022-02-14 | 22 | 43 |  | 600 |
| 2022-02-21 | 17 | 40 |  | 600 |

Nunber of infections

| Week | Primary infections | Delta | Omicron |
| --- | --- | --- | --- |
| 2021-09-06 | 1 | 600 |  |
| 2021-09-13 | 0 | 600 |  |
| 2021-09-20 | 6 | 600 |  |
| 2021-09-27 | 4 | 600 |  |
| 2021-10-04 | 15 | 600 |  |
| 2021-10-11 | 11 | 600 |  |
| 2021-10-18 | 8 | 600 |  |
| 2021-10-25 | 14 | 600 |  |
| 2021-11-01 | 10 | 600 |  |
| 2021-11-08 | 19 | 600 |  |
| 2021-11-15 | 31 | 600 |  |
| 2021-11-22 | 31 | 600 |  |
| 2021-11-29 | 44 |  |  |
| 2021-12-06 | 57 |  |  |
| 2021-12-13 | 149 |  | 600 |
| 2021-12-20 | 246 |  | 600 |
| 2021-12-27 | 509 |  | 600 |
| 2022-01-03 | 385 |  | 600 |
| 2022-01-10 | 189 |  | 600 |
| 2022-01-17 | 189 |  | 600 |
| 2022-01-24 | 246 |  | 600 |
| 2022-01-31 | 201 |  | 600 |
| 2022-02-07 | 162 |  | 600 |
| 2022-02-14 | 119 |  | 600 |
| 2022-02-21 | 100 |  | 600 |

Footnote: During the Delta analysis period (07 September 2021 to 30 November 2021) in the previously infected cohort: primary infections n=367; reinfection n=23 and in the previously uninfected cohort: primary infections n=194. During the Omicron analysis period (13 December 2021 to 28 February 2022) in the previously infected cohort: primary infections n=318; reinfection n=897 and in the previously uninfected cohort: primary infections n=2,498.

### **List of abbreviations used in tables**

- HR: hazard ratio.
- CI: confidence interval.
- PF: Pfizer (BNT162b2) vaccine.
- AZ: AstraZeneca (ChAdOx1) vaccine.
- d2: two vaccine doses.
- d2 194-253: between 194 and 353 days after dose 2 (example).
- d2 254+: 254 or more days after dose 2.
- d3: three vaccine doses.
- d3 7-66: between 7 and 7 days after dose 3 (example).
- 3PF: three doses of Pfizer vaccine (primary course + booster).
- 2AZ/1PF: primary course AstraZeneca, 1 Pfizer booster.
- PI: previous infection
- PI *<* 1yr: previous infection less than one year ago (example).

**Statistical Appendix A**

### Statistical Appendix A: Full model outputs

We report results for the adjusted models presented in the main text, here including the estimates of hazard ratios (HR) for additional predictors. We report only HR (not vaccine effectiveness) for all predictors.

#### A.1 Booster effectiveness in negative cohort

##### A.1.1 Delta period

Table A.1: Full output for adjusted model that estimates booster (V3) effec- tiveness in negative cohort (Table 1 of main text), during Delta period. The estimates of some coefficients did not converge (NC).

|  | | **HR** | **95% CI** |
| --- | --- | --- | --- |
| **Vaccination status**  d3 0-6 3PF |  | 0.65 | (0.33-1.27) |
| d3 7+ 3PF |  | 0.37 | (0.23-0.60) |
| d2 194+ AZ |  | 1.58 | (0.76-3.28) |
| d3 0-6 2AZ/1PF |  | 1.91 | (0.69-5.30) |
| d3 7+ 2AZ/1PF |  | 0.44 | (0.18-1.08) |
| **Gender**  Female |  | 0.62 | (0.47-0.83) |
| Other |  | NC | - |
| **Ethnic group**  Asian |  | 0.84 | (0.50-1.41) |
| Black |  | 0.56 | (0.14-2.25) |
| Mixed |  | 0.75 | (0.26-2.19) |
| Other |  | 0.44 | (0.11-1.72) |
|  | 5 |  |  |

**Table A.1 continued from previous page**

|  | **HR** | **95% CI** |
| --- | --- | --- |
| Prefer not to say | NC | - |
| **Staff type**  Doctor | 1.37 | (0.84-2.23) |
| Nursing | 1.29 | (0.81-2.03) |
| Healthcare Assistant | 1.79 | (1.03-3.11) |
| Midwife | 1.80 | (0.76-4.27) |
| Healthcare Scientist | 1.69 | (0.94-3.03) |
| Pharmacist | 1.00 | (0.48-2.06) |
| Physiotherapist/Occupational Therapist/SALT | 1.09 | (0.58-2.05) |
| Student (Medical/Nursing/Midwifery/Other) | 1.30 | (0.48-3.48) |
| Estates/Porters/Security | 1.62 | (0.72-3.61) |
| Other - no patient contact | 1.43 | (0.90-2.28) |
| Other - patient contact | 1.15 | (0.44-3.04) |

A.1. *BOOSTER EFFECTIVENESS IN NEGATIVE COHORT*

##### A.1.2 Omicron period

Table A.2: Full output for adjusted model that estimates booster (V3) effec- tiveness in negative cohort (Table 1 of main text), during Omicron period.

|  | **HR** | **95% CI** |
| --- | --- | --- |
| **Vaccination status**  d3 0-6 3PF | 1.13 | (0.65-1.97) |
| d3 7-66 3PF | 0.65 | (0.53-0.79) |
| d3 67-126 3PF | 0.66 | (0.57-0.78) |
| d3 127+ 3PF | 0.79 | (0.62-1) |
| d2 194+ AZ | 1.22 | (0.81-1.84) |
| d3 7-66 2AZ/1PF | 0.39 | (0.27-0.56) |
| d3 67+ 2AZ/1PF | 0.54 | (0.40-0.72) |
| **Gender**  Female | 1.13 | (1-1.28) |
| Other | 0.98 | (0.36-2.69) |
| **Ethnic group**  Asian | 1.11 | (0.92-1.33) |
| Black | 1.17 | (0.83-1.65) |
| Mixed | 0.97 | (0.68-1.39) |
| Other | 1.24 | (0.86-1.79) |
| Prefer not to say | 1.53 | (0.68-3.43) |
| **Staff type**  Doctor | 0.70 | (0.59-0.83) |
| Nursing | 0.80 | (0.72-0.90) |
| Healthcare Assistant | 1.12 | (0.94-1.34) |
| Midwife | 0.89 | (0.66-1.21) |
| Healthcare Scientist | 0.72 | (0.58-0.89) |
| Pharmacist | 0.64 | (0.47-0.87) |
| Physiotherapist/Occupational Therapist/SALT | 0.87 | (0.69-1.11) |
| Student (Medical/Nursing/Midwifery/Other) | 0.92 | (0.70-1.19) |
| Estates/Porters/Security | 1.28 | (0.92-1.78) |
| Other - no patient contact | 0.77 | (0.65-0.91) |
| Other - patient contact | 0.75 | (0.54-1.05) |

#### A.2 Protection associated with primary infec tion

##### A.2.1 Delta period

Table A.3: Full output for adjusted model that estimates protection from reinfection associated with primary infection (Table 2 of main text), during Delta period. The estimates of some coefficients did not converge (NC).

|  | **HR** | **95% CI** |
| --- | --- | --- |
| **Primary infection status**  3-6 mth ago | 0.10 | (0.02-0.58) |
| 6-9 mth ago | 0.25 | (0.15-0.41) |
| 9-12 mth ago | 0.37 | (0.26-0.52) |
| 12-15 mth ago | 0.07 | (0.02-0.28) |
| 15+ mth ago | 0.15 | (0.11-0.19) |
| **Vaccination status**  d2 194-253 PF | 1.33 | (1.08-1.65) |
| d2 254+ PF | 1.48 | (0.99-2.21) |
| d3 0-6 3PF | 1.04 | (0.69-1.56) |
| d3 7+ 3PF | 0.57 | (0.43-0.77) |
| d2 194+ AZ | 2.42 | (1.43-4.10) |
| d3 0-6 2AZ/1PF | 2.81 | (1.19-6.67) |
| d3 7+ 2AZ/1PF | 0.73 | (0.32-1.67) |
| **Gender**  Female | 0.71 | (0.61-0.84) |
| Other | NC | - |
| **Ethnic group**  Asian | 0.95 | (0.72-1.25) |
| Black | 1.03 | (0.60-1.76) |
| Mixed | 0.94 | (0.58-1.53) |
| Other | 0.67 | (0.37-1.23) |
| Prefer not to say | 0.60 | (0.08-4.47) |
| **Staff type**  Doctor | 1.02 | (0.81-1.29) |
| Nursing | 1.06 | (0.86-1.31) |
| Healthcare Assistant | 1.36 | (1.06-1.75) |
| Midwife | 1.20 | (0.71-2.02) |
| Healthcare Scientist | 1.18 | (0.91-1.53) |
| Pharmacist | 0.93 | (0.64-1.33) |

A.2. *PROTECTION ASSOCIATED WITH PRIMARY INFECTION*

**Table A.3 continued from previous page**

|  | **HR** | **95% CI** |
| --- | --- | --- |
| Physiotherapist/Occupational Therapist/SALT | 1.14 | (0.84-1.56) |
| Student (Medical/Nursing/Midwifery/Other) | 1.14 | (0.75-1.72) |
| Estates/Porters/Security | 0.98 | (0.63-1.51) |
| Other - no patient contact | 1.08 | (0.83-1.40) |
| Other - patient contact | 1.26 | (0.75-2.10) |

##### A.2.2 Omicron period

Table A.4: Full output for adjusted model that estimates protection from re infection associated with primary infection (Table 2 of main text), during the Omicron period.

|  | **HR** | **95% CI** |
| --- | --- | --- |
| **Vaccination status**  d3 0-6 3PF | 0.93 | (0.55-1.59) |
| d3 7-66 3PF | 0.63 | (0.53-0.74) |
| d3 67-126 3PF | 0.67 | (0.59-0.77) |
| d3 127+ 3PF | 0.81 | (0.66-0.99) |
| d2 194+ AZ | 1.08 | (0.81-1.44) |
| d3 0-6 2AZ/1PF | 0.42 | (0.11-1.64) |
| d3 7-66 2AZ/1PF | 0.45 | (0.35-0.59) |
| d3 67+ 2AZ/1PF | 0.51 | (0.40-0.64) |
| **Primary infection status**  3-6 mth ago | 0.33 | (0.25-0.44) |
| 6-9 mth ago | 0.47 | (0.27-0.80) |
| 9-12 mth ago | 0.73 | (0.56-0.96) |
| 12-15 mth ago | 0.73 | (0.63-0.84) |
| 15+ mth ago | 0.74 | (0.66-0.83) |
| **Gender**  Female | 1.12 | (1-1.27) |
| Other | 0.84 | (0.32-2.21) |
| **Ethnic group**  Asian | 1.12 | (0.95-1.31) |
| Black | 1.18 | (0.88-1.57) |
| Mixed | 1.08 | (0.81-1.44) |
| Other | 1.20 | (0.90-1.61) |
| Prefer not to say | 1.37 | (0.61-3.08) |

**Table A.4 continued from previous page**

|  | **HR** | **95% CI** |
| --- | --- | --- |
| **Staff type** |  |  |
| Doctor | 0.75 | (0.64-0.87) |
| Nursing | 0.85 | (0.76-0.94) |
| Healthcare Assistant | 1.04 | (0.89-1.21) |
| Midwife | 0.84 | (0.64-1.10) |
| Healthcare Scientist | 0.77 | (0.62-0.95) |
| Pharmacist | 0.65 | (0.51-0.85) |
| Physiotherapist/Occupational Therapist/SALT | 0.84 | (0.68-1.04) |
| Student (Medical/Nursing/Midwifery/Other) | 0.92 | (0.74-1.14) |
| Estates/Porters/Security | 1.14 | (0.86-1.51) |
| Other - no patient contact | 0.76 | (0.65-0.89) |
| Other - patient contact | 0.83 | (0.61-1.11) |

#### A.3 Hybrid protection

##### A.3.1 Delta period

Table A.5: Full output for adjusted model that estimates hybrid protection (Table 3 of main text), during Delta period. The estimates of numerous coefficients did not converge (NC).

|  | **HR** | **95% CI** |
| --- | --- | --- |
| **Vaccination status**  d3 0-6 3PF PI *<*=1yr ago | 0.19 | (0.02-2.11) |
| d3 7+ 3PF PI *<*=1yr ago | 0.12 | (0.04-0.34) |
| d3 0-6 3PF PI *>*1yr ago | 0.22 | (0.04-1.06) |
| d3 7+ 3PF PI *>*1yr ago | 0.10 | (0.04-0.24) |
| **Gender**  Female | 1.38 | (0.55-3.43) |
| Other | NC | - |
| **Ethnic group**  Asian | 0.24 | (0.02-2.36) |
| Black | NC | - |
| Mixed | NC | - |
| Other | NC | - |
| Prefer not to say | NC | - |
| **Staff type**  Doctor | 0.76 | (0.17-3.42) |

A.3. *HYBRID PROTECTION*

**Table A.5 continued from previous page**

|  | **HR** | **95% CI** |
| --- | --- | --- |
| Nursing | 0.83 | (0.21-3.27) |
| Healthcare Assistant | 0.58 | (0.06-5.46) |
| Midwife | NC | - |
| Healthcare Scientist | NC | - |
| Pharmacist | 0.88 | (0.10-8.15) |
| Physiotherapist/Occupational Therapist/SALT | 0.25 | (0.02-2.66) |
| Student (Medical/Nursing/Midwifery/Other) | 0.96 | (0.08-10.87) |
| Estates/Porters/Security | 2.46 | (0.48-12.68) |
| Other - no patient contact | 0.50 | (0.12-2.13) |
| Other - patient contact | NC | - |

##### A.3.2 Omicron period

Table A.6: Full output for adjusted model that estimates hybrid protection (Table 3 of main text), during Omicron period. The estimates of one coefficient did not converge (NC).

|  | **HR** | **95% CI** |
| --- | --- | --- |
| **Vaccination status**  d2 194-253 PF PI *<*=1yr | 0.79 | (0.38-1.67) |
| d2 254+ PF PI *<*=1yr | 0.45 | (0.20-0.99) |
| d3 0-6 3PF PI *<*=1yr | 0.59 | (0.16-2.13) |
| d3 7-66 3PF PI *<*=1yr | 0.34 | (0.22-0.51) |
| d3 67-126 3PF PI *<*=1yr | 0.32 | (0.24-0.43) |
| d3 127+ 3PF PI *<*=1yr | 0.23 | (0.08-0.67) |
| d2 194-253 PF PI *>*1yr | 0.83 | (0.46-1.48) |
| d2 254+ PF PI *>*1yr | 0.68 | (0.50-0.91) |
| d3 0-6 3PF PI *>*1yr | 0.44 | (0.10-1.87) |
| d3 7-66 3PF PI *>*1yr | 0.43 | (0.34-0.55) |
| d3 67-126 3PF PI *>*1yr | 0.52 | (0.43-0.62) |
| d3 127+ 3PF PI *>*1yr | 0.59 | (0.42-0.83) |
| **Gender**  Female | 1.09 | (0.85-1.39) |
| Other | NC | - |
| **Ethnic group**  Asian | 1.16 | (0.93-1.43) |
| Black | 1.33 | (0.83-2.15) |
| Mixed | 1.16 | (0.73-1.83) |
| Other | 1.07 | (0.72-1.58) |

**Table A.6 continued from previous page**

|  | **HR** | **95% CI** |
| --- | --- | --- |
| Prefer not to say | 0.66 | (0.07-6.40) |
| **Staff type**  Doctor | 0.71 | (0.56-0.89) |
| Nursing | 0.86 | (0.70-1.06) |
| Healthcare Assistant | 0.89 | (0.65-1.21) |
| Midwife | 0.92 | (0.57-1.47) |
| Healthcare Scientist | 0.88 | (0.59-1.30) |
| Pharmacist | 0.75 | (0.46-1.22) |
| Physiotherapist/Occupational Therapist/SALT | 0.74 | (0.52-1.04) |
| Student (Medical/Nursing/Midwifery/Other) | 0.89 | (0.56-1.39) |
| Estates/Porters/Security | 0.98 | (0.61-1.59) |
| Other - no patient contact | 0.69 | (0.56-0.84) |
| Other - patient contact | 1.08 | (0.57-2.04) |

**Statistical Appendix B**

### Statistical Appendix B: Sensitivity analyses for depletion-of-susceptible bias

#### B.1 Delta period

We ran a sensitivity analysis to investigate if the depletion of susceptible bias affected our estimates of protection against the Delta variant.

The only model to which this analysis applies is the model estimating protection from primary infection. Models investigating booster’s vaccine effectiveness do not have time-dependent, as only one time interval was considered for lack of follow-up time, the interval being approximately 0-2 months after booster. As no waning can be identified with a single time-interval, the depletion of susceptible bias cannot be ascertained.

For the model estimating protection from primary infection, we stopped the study at November 1st 2021, instead of December 1st 2021. For the estimates of protection from primary infection, results are more uncertain, but compatible with those obtained considering the full dataset. We conclude that there is no evidence of the depletion of susceptible bias in this dataset. The estimates of some HRs did not converge, flagged by NC.

Table B.1: Results of sensitivity analysis for depletion-of-susceptible bias in Delta period, for the model estimating protection from primary infection. The estimates of some HRs did not converge, flagged by NC.

|  | **HR** | **95% CI** |
| --- | --- | --- |
| **Primary infection status** |  |  |
| 3-6 mth ago | 0.18 | (0.03-1.01) |
| 6-9 mth ago | 0.25 | (0.15-0.42) |
| 9-12 mth ago | 0.42 | (0.29-0.61) |
| 12-15 mth ago | NC | - |

**Table B.1 continued from previous page**

|  | **HR** | **95% CI** |
| --- | --- | --- |
| 15+ mth ago | 0.13 | (0.09-0.18) |
| **Vaccine status**  d2 134-193 PF | Ref. |  |
| d2 194-253 PF | 1.36 | (1.10-1.69) |
| d2 254+ PF | 1.24 | (0.73-2.11) |
| d3 0-6 3PF | 1.04 | (0.70-1.54) |
| d3 7+ 3PF | 0.67 | (0.46-0.98) |
| d2 194+ AZ | 1.98 | (0.76-5.11) |
| d3 0-6 2AZ/1PF | 2.53 | (0.95-6.75) |
| d3 7+ 2AZ/1PF | 1.08 | (0.27-4.42) |
| **Gender**  Female | 0.75 | (0.63-0.90) |
| Other | NC | - |
| **Ethnic group**  Asian | 1.03 | (0.76-1.39) |
| Black | 0.98 | (0.52-1.85) |
| Mixed | 1.00 | (0.60-1.69) |
| Other | 0.77 | (0.40-1.46) |
| Prefer not to say | 0.75 | (0.10-5.54) |
| **Staff type**  Doctor | 0.93 | (0.72-1.20) |
| Nursing | 1.00 | (0.79-1.26) |
| Healthcare Assistant | 1.24 | (0.94-1.63) |
| Midwife | 1.22 | (0.71-2.09) |
| Healthcare Scientist | 1.07 | (0.81-1.43) |
| Pharmacist | 0.87 | (0.58-1.31) |
| Physiotherapist/Occupational Therapist/SALT | 1.11 | (0.79-1.56) |
| Student (Medical/Nursing/Midwifery/Other) | 1.13 | (0.70-1.82) |
| Estates/Porters/Security | 0.86 | (0.51-1.44) |
| Other- patient contact | 1.00 | (0.75-1.35) |
| Other - no patient contact | 1.31 | (0.76-2.28) |

#### B.2 Omicron period

In order to investigate the depletion-of-susceptible bias in the Omicron period, we stopped the study at January 31, 2022, instead of February 28, 2022. Here the sensitivity analysis applies to all models: naive protection, hybrid protection and protection from primary infection. Results are more uncertain, but

compatible with those obtained considering the full dataset. We conclude that there is no evidence of the depletion of susceptible bias.

Table B.2: Sensitivity-analysis for effectiveness of booster (V3) in negative co- hort, Omicron period.

|  | **HR** | **95% CI** |
| --- | --- | --- |
| **Vaccine status**  d3 0-6 3PF | 1.19 | (0.67-2.11) |
| d3 7-66 3PF | 0.67 | (0.55-0.82) |
| d3 67-126 3PF | 0.67 | (0.57-0.79) |
| d3 127+ 3PF | 1.13 | (0.64-2) |
| d2 194+ AZ | 1.36 | (0.88-2.09) |
| d3 7-66 2AZ/1PF | 0.38 | (0.26-0.56) |
| d3 67+ 2AZ/1PF | 0.57 | (0.41-0.80) |
| **Gender**  Female | 1.14 | (0.98-1.32) |
| Other | 0.35 | (0.06-2.22) |
| **Ethnic group**  Asian | 1.04 | (0.83-1.31) |
| Black | 1.19 | (0.83-1.71) |
| Mixed | 0.73 | (0.45-1.21) |
| Other | 1.28 | (0.81-2.03) |
| Prefer not to say | 1.39 | (0.56-3.46) |
| **Staff type**  Doctor | 0.68 | (0.55-0.84) |
| Nursing | 0.77 | (0.68-0.88) |
| Healthcare Assistant | 1.08 | (0.89-1.32) |
| Midwife | 1.00 | (0.71-1.41) |
| Healthcare Scientist | 0.76 | (0.60-0.96) |
| Pharmacist | 0.63 | (0.45-0.87) |
| Physiotherapist/Occupational Therapist/SALT | 0.79 | (0.60-1.04) |
| Student (Medical/Nursing/Midwifery/Other) | 0.78 | (0.55-1.10) |
| Estates/Porters/Security | 1.21 | (0.87-1.69) |
| Other - no patient contact | 0.74 | (0.61-0.91) |
| Other - patient contact | 0.87 | (0.61-1.23) |

Table B.3: Sensitivity-analysis for hybrid protection, Omicron period.

|  | **HR** | **95% CI** |
| --- | --- | --- |
| **Vaccine status**  d2 194-253 PF PI *<*=1yr | 0.78 | (0.38-1.61) |
| d2 254+ PF PI *<*=1yr | 0.48 | (0.22-1.07) |
| d3 0-6 3PF PI *<*=1yr | 0.62 | (0.17-2.21) |
| d3 7-66 3PF PI *<*=1yr | 0.31 | (0.21-0.48) |
| d3 67-126 3PF PI *<*=1yr | 0.31 | (0.22-0.43) |
| d3 127+ 3PF PI *<*=1yr | NC | - |
| d2 194-253 PF PI *>*1yr | 0.88 | (0.46-1.69) |
| d2 254+ PF PI *>*1yr | 0.70 | (0.51-0.96) |
| d3 0-6 3PF PI *>*1yr | 0.44 | (0.10-1.90) |
| d3 7-66 3PF PI *>*1yr | 0.41 | (0.32-0.53) |
| d3 67-126 3PF PI *>*1yr | 0.51 | (0.42-0.63) |
| d3 127+ 3PF PI *>*1yr | 0.87 | (0.31-2.44) |
| **Gender**  Female | 1.13 | (0.86-1.50) |
| Other | NC | - |
| **Ethnic group**  Asian | 1.14 | (0.89-1.47) |
| Black | 1.19 | (0.72-1.96) |
| Mixed | 1.27 | (0.78-2.07) |
| Other | 1.18 | (0.78-1.77) |
| Prefer not to say | 0.78 | (0.08-8) |
| **Staff type**  Doctor | 0.64 | (0.50-0.82) |
| Nursing | 0.78 | (0.63-0.96) |
| Healthcare Assistant | 0.95 | (0.69-1.31) |
| Midwife | 0.99 | (0.62-1.58) |
| Healthcare Scientist | 0.77 | (0.50-1.17) |
| Pharmacist | 0.82 | (0.52-1.29) |
| Physiotherapist/Occupational Therapist/SALT | 0.70 | (0.49-1.01) |
| Student (Medical/Nursing/Midwifery/Other) | 0.82 | (0.50-1.35) |
| Estates/Porters/Security | 0.94 | (0.60-1.48) |
| Other - no patient contact | 0.64 | (0.51-0.80) |
| Other - patient contact | 1.09 | (0.54-2.19) |

Protection associated with primary infection:

Table B.4: Sensitivity-analysis for protection from primary infection, Omicron period.

|  | **HR** | **95% CI** |
| --- | --- | --- |
| **Primary infection status**  3-6 mth ago | 0.28 | (0.20-0.40) |
| 6-9 mth ago | 0.67 | (0.37-1.20) |
| 9-12 mth ago | 0.72 | (0.54-0.95) |
| 12-15 mth ago | 0.70 | (0.61-0.81) |
| 15+ mth ago | 0.76 | (0.66-0.86) |
| **Vaccination status**  d3 0-6 3PF | 0.97 | (0.57-1.66) |
| d3 7-66 3PF | 0.63 | (0.53-0.75) |
| d3 67-126 3PF | 0.68 | (0.59-0.78) |
| d3 127+ 3PF | 1.12 | (0.68-1.84) |
| d2 194+ AZ | 1.15 | (0.86-1.54) |
| d3 0-6 2AZ/1PF | 0.43 | (0.11-1.69) |
| d3 7-66 2AZ/1PF | 0.43 | (0.33-0.55) |
| d3 67+ 2AZ/1PF | 0.53 | (0.42-0.68) |
| **Ethnic group**  Asian | 1.08 | (0.87-1.33) |
| Black | 1.14 | (0.84-1.54) |
| Mixed | 0.94 | (0.67-1.33) |
| Other | 1.30 | (0.90-1.86) |
| Prefer not to say | 1.29 | (0.52-3.22) |
| **Staff type**  Doctor | 0.71 | (0.58-0.85) |
| Nursing | 0.81 | (0.71-0.91) |
| Healthcare Assistant | 1.04 | (0.88-1.23) |
| Midwife | 0.91 | (0.68-1.21) |
| Healthcare Scientist | 0.77 | (0.61-0.96) |
| Pharmacist | 0.64 | (0.49-0.85) |
| Physiotherapist/Occupational Therapist/SALT | 0.79 | (0.62-1.01) |
| Student (Medical/Nursing/Midwifery/Other) | 0.83 | (0.62-1.11) |
| Estates/Porters/Security | 1.06 | (0.79-1.42) |
| Other - no patient contact | 0.73 | (0.61-0.88) |
| Other - patient contact | 0.94 | (0.68-1.29) |

**Statistical Appendix C**

### Statistical Appendix C: Sensitivity analyses for delayed-entry bias

We ran sensitivity analyses to assess the effect of having delayed entry in our studies.

#### C.1 Delta period

For the Delta period, we reran the models including only participants who did not enroll in the second-year extension, i.e. without delayed-entry.

Table C.1: Results of sensitivity analyses for delayed-entry bias, for model estimating effectiveness of booster in negative cohort during Delta period. The estimates of some HRs did not converge (NC).

|  | **HR** | **95 % CI** |
| --- | --- | --- |
| **Vaccine status**  d3 0-6 3PF | 0.79 | (0.36-1.70) |
| d3 7+ 3PF | 0.41 | (0.23-0.73) |
| d2 194+ AZ | 1.71 | (0.73-4.01) |
| d3 0-6 2AZ/1PF | 2.73 | (0.45-16.61) |
| d3 7+ 2AZ/1PF | 0.40 | (0.08-1.95) |
| **Gender**  Female | 0.66 | (0.42-1.04) |
| Other | NC | - |
| **Ethnic group**  Asian | 1.10 | (0.57-2.13) |
| Black | 0.80 | (0.23-2.77) |

**Table C.1 continued from previous page**

|  | **HR** | **95 % CI** |
| --- | --- | --- |
| Mixed | 1.35 | (0.43-4.27) |
| Other | 0.44 | (0.05-3.87) |
| Prefer not to say | NC | - |
| **Staff type**  Doctor | 1.58 | (0.77-3.23) |
| Nursing | 1.88 | (0.96-3.69) |
| Healthcare Assistant | 1.89 | (0.76-4.71) |
| Midwife | 1.36 | (0.20-9.05) |
| Healthcare Scientist | 2.54 | (1.25-5.18) |
| Pharmacist | 0.91 | (0.35-2.41) |
| Physiotherapist/Occupational Therapist/SALT | 1.88 | (0.82-4.32) |
| Student (Medical/Nursing/Midwifery/Other) | NC | - |
| Estates/Porters/Security | 1.47 | (0.35-6.12) |
| Other - no patient contact | 2.18 | (1.05-4.53) |
| Other - patient contact | 2.95 | (0.77-11.28) |

Table C.2: Results of sensitivity analyses for delayed-entry bias, for model esti- mating hybrid protection during Delta period. The estimates of numerous HRs did not converge (NC).

|  | **HR** | **95% CI** |
| --- | --- | --- |
| **Vaccine and PI status**  d3 0-6 3PF PI *<*=1yr ago | NC | - |
| d3 7+ 3PF PI *<*=1yr ago | 0.08 | (0.01-0.69) |
| d3 0-6 3PF PI *>*1yr ago | NC | - |
| d3 7+ 3PF PI *>*1yr ago | 0.06 | (0.01-0.31) |
| **Gender**  Female | 1.11 | (0.16-7.69) |
| Other | NC | - |
| **Ethnic group**  Asian | 0.98 | (0.11-8.84) |
| Black | NC | - |
| Mixed | NC | - |
| Other | NC | - |
| Prefer | NC | - |
| **Staff type**  Doctor | 0.37 | (0.05-2.92) |

*C.1. DELTA PERIOD*

**Table C.2 continued from previous page**

|  | **HR** | **95% CI** |
| --- | --- | --- |
| Nursing | 0.79 | (0.22-2.87) |
| Healthcare Assistant | 0.51 | (0.05-5.69) |
| Midwife | NC | - |
| Healthcare Scientist | NC | - |
| Pharmacist | NC | - |
| Physiotherapist/Occupational Therapist/SALT | 0.47 | (0.03-6.40) |
| Student (Medical/Nursing/Midwifery/Other) | NC | - |
| Estates/Porters/Security | NC | - |
| Other - no patient contact | 0.50 | (0.10-2.59) |
| Other - patient contact | NC | - |

Table C.3: Results of sensitivity analyses for delayed-entry bias, for model esti- mating protection associated with primary infection during Delta period. The estimates of some HRs did not converge (NC).

|  | **HR** | **95% CI** |
| --- | --- | --- |
| **Primary infection status**  3-6 mth ago | 0.17 | (0.03-0.89) |
| 6-9 mth ago | 0.29 | (0.14-0.59) |
| 9-12 mth ago | 0.38 | (0.23-0.63) |
| 12-15 mth ago | NC | - |
| 15+ mth ago | 0.14 | (0.08-0.23) |
| **Vaccination status**  d2 194-253 PF | 1.29 | (0.96-1.72) |
| d2 254+ PF | 1.59 | (1-2.53) |
| d3 0-6 3PF | 1.28 | (0.71-2.30) |
| d3 7+ 3PF | 0.67 | (0.44-1.03) |
| d2 194+ AZ | 2.56 | (1.23-5.33) |
| d3 0-6 2AZ/1PF | 3.87 | (0.77-19.37) |
| d3 7+ 2AZ/1PF | 0.72 | (0.17-3) |
| **Gender**  Female | 0.62 | (0.51-0.75) |
| Other | NC | - |
| **Ethnic group**  Asian | 1.00 | (0.71-1.41) |
| Black | 1.09 | (0.58-2.07) |
| Mixed | 1.01 | (0.52-1.97) |
| Other | 0.79 | (0.37-1.69) |

**Table C.3 continued from previous page**

|  | **HR** | **95% CI** |
| --- | --- | --- |
| Prefer not to say | 0.80 | (0.11-5.90) |
| **Staff type**  Doctor | 0.95 | (0.71-1.27) |
| Nursing | 1.17 | (0.89-1.55) |
| Healthcare Assistant | 1.47 | (1.06-2.02) |
| Midwife | 0.81 | (0.36-1.85) |
| Healthcare Scientist | 1.24 | (0.93-1.65) |
| Pharmacist | 0.88 | (0.51-1.52) |
| Physiotherapist/Occupational Therapist/SALT | 1.01 | (0.65-1.58) |
| Student (Medical/Nursing/Midwifery/Other) | 1.54 | (0.79-3.02) |
| Estates/Porters/Security | 0.95 | (0.53-1.71) |
| Other - no patient contact | 1.17 | (0.81-1.68) |
| Other - patient contact | 1.26 | (0.54-2.92) |
| **C.2 Omicron period** |  |  |

For the Omicron period, we reran the models including only participants who *did* enroll in the second-year extension. This is an alternative way to assess the potential impact of delayed-entry bias. An analysis with only participants who did not enroll in the extension has been attempted as well, but has been found to be severely under powered. The reason is that almost all participants who were in SIREN during the Omicron period were those who had enrolled in the second-year extension.

Table C.4: Results of sensitivity analyses (including only participants who en- rolled in the extension) for model estimating effectiveness of booster in negative cohort during Omicron period.

|  | **HR** | **95% Ci** |
| --- | --- | --- |
| **Vaccination status**  d3 0-6 3PF | 1.13 | (0.48-2.69) |
| d3 7-66 3PF | 0.67 | (0.48-0.93) |
| d3 67-126 3PF | 0.70 | (0.53-0.94) |
| d3 127+ 3PF | 0.85 | (0.59-1.21) |
| d2 194+ AZ | 1.46 | (0.81-2.62) |
| d3 7-66 2AZ/1PF | 0.42 | (0.25-0.68) |
| d3 67+ 2AZ/1PF | 0.62 | (0.42-0.91) |
| **Gender**  Female | 1.18 | (1-1.39) |
| Other | 1.26 | (0.45-3.48) |

**Table C.4 continued from previous page**

|  | **HR** | **95% Ci** |
| --- | --- | --- |
| **Ethnic group**  Asian | 1.21 | (0.98-1.48) |
| Black | 1.08 | (0.62-1.85) |
| Mixed | 1.09 | (0.72-1.66) |
| Other | 1.30 | (0.78-2.19) |
| Prefer not to say | 1.59 | (0.43-5.90) |
| **Staff type**  Doctor | 0.72 | (0.57-0.90) |
| Nursing | 0.82 | (0.70-0.96) |
| Healthcare Assistant | 1.07 | (0.84-1.37) |
| Midwife | 0.90 | (0.64-1.26) |
| Healthcare Scientist | 0.81 | (0.61-1.08) |
| Pharmacist | 0.62 | (0.40-0.95) |
| Physiotherapist/Occupational Therapist/SALT | 0.81 | (0.60-1.11) |
| Student (Medical/Nursing/Midwifery/Other) | 1.02 | (0.76-1.36) |
| Estates/Porters/Security | 1.40 | (0.95-2.08) |
| Other - no patient contact | 0.80 | (0.65-1) |
| Other - patient contact | 0.86 | (0.58-1.26) |

Table C.5: Results of sensitivity analyses (including only participants who en- rolled in the extension) for model estimating hybrid protection during Omicron period. The estimates of some coefficients did not converge (NC).

|  | **HR** | **95% CI** |
| --- | --- | --- |
| **Vaccination and PI status**  d2 194-253 PF PI *<*=1yr | 0.79 | (0.38-1.67) |
| d2 254+ PF PI *<*=1yr | 0.45 | (0.20-0.99) |
| d3 0-6 3PF PI *<*=1yr | 0.59 | (0.16-2.13) |
| d3 7-66 3PF PI *<*=1yr | 0.34 | (0.22-0.51) |
| d3 67-126 3PF PI *<*=1yr | 0.32 | (0.24-0.43) |
| d3 127+ 3PF PI *<*=1yr | 0.23 | (0.08-0.67) |
| d2 194-253 PF PI *>*1yr | 0.83 | (0.46-1.48) |
| d2 254+ PF PI *>*1yr | 0.68 | (0.50-0.91) |
| d3 0-6 3PF PI *>*1yr | 0.44 | (0.10-1.87) |
| d3 7-66 3PF PI *>*1yr | 0.43 | (0.34-0.55) |
| d3 67-126 3PF PI *>*1yr | 0.52 | (0.43-0.62) |
| d3 127+ 3PF PI *>*1yr | 0.59 | (0.42-0.83) |
| **Gender** |  |  |

**Table C.5 continued from previous page**

|  | **HR** | **95% CI** |
| --- | --- | --- |
| Female | 1.09 | (0.85-1.39) |
| Other | NC | - |
| **Ethnic group**  Asian | 1.16 | (0.93-1.43) |
| Black | 1.33 | (0.83-2.15) |
| Mixed | 1.16 | (0.73-1.83) |
| Other | 1.07 | (0.72-1.58) |
| Prefer not to say | 0.66 | (0.07-6.40) |
| **Staff type**  Doctor | 0.71 | (0.56-0.89) |
| Nursing | 0.86 | (0.70-1.06) |
| Healthcare Assistant | 0.89 | (0.65-1.21) |
| Midwife | 0.92 | (0.57-1.47) |
| Healthcare Scientist | 0.88 | (0.59-1.30) |
| Pharmacist | 0.75 | (0.46-1.22) |
| Physiotherapist/Occupational Therapist/SALT | 0.74 | (0.52-1.04) |
| Student (Medical/Nursing/Midwifery/Other) | 0.89 | (0.56-1.39) |
| Estates/Porters/Security | 0.98 | (0.61-1.59) |
| Other - no patient contact | 0.69 | (0.56-0.84) |
| Other - patient contact | 1.08 | (0.57-2.04) |

Table C.6: Results of sensitivity analyses (including only participants who en- rolled in the extension) for model estimating protection associated with primary infection during Omicron period.

|  | **HR** | **95% CI** |
| --- | --- | --- |
| **Primary Infection Status**  3-6 mth ago | 0.36 | (0.26-0.52) |
| 6-9 mth ago | 0.47 | (0.25-0.87) |
| 9-12 mth ago | 0.73 | (0.54-1) |
| 12-15 mth ago | 0.82 | (0.69-0.98) |
| 15+ mth ago | 0.80 | (0.71-0.91) |
| **Vaccination status**  d3 0-6 3PF | 0.83 | (0.41-1.66) |
| d3 7-66 3PF | 0.64 | (0.49-0.83) |
| d3 67-126 3PF | 0.70 | (0.56-0.88) |
| d3 127+ 3PF | 0.82 | (0.62-1.09) |
| d2 194+ AZ | 1.06 | (0.73-1.53) |

**Table C.6 continued from previous page**

|  | **HR** | **95% CI** |
| --- | --- | --- |
| d3 0-6 2AZ/1PF | 0.33 | (0.04-2.72) |
| d3 7-66 2AZ/1PF | 0.46 | (0.33-0.65) |
| d3 67+ 2AZ/1PF | 0.54 | (0.40-0.73) |
| **Gender**  Female | 1.18 | (1.01-1.37) |
| Other | 0.95 | (0.36-2.51) |
| **Ethnic group**  Asian | 1.20 | (0.99-1.46) |
| Black | 1.07 | (0.73-1.57) |
| Mixed | 1.22 | (0.89-1.66) |
| Other | 1.21 | (0.83-1.77) |
| Prefer not to say | 1.09 | (0.30-3.95) |
| **Staff type**  Doctor | 0.79 | (0.66-0.95) |
| Nursing | 0.85 | (0.74-0.97) |
| Healthcare Assistant | 0.94 | (0.77-1.16) |
| Midwife | 0.76 | (0.56-1.03) |
| Healthcare Scientist | 0.82 | (0.62-1.07) |
| Pharmacist | 0.66 | (0.47-0.92) |
| Physiotherapist/Occupational Therapist/SALT | 0.79 | (0.60-1.04) |
| Student (Medical/Nursing/Midwifery/Other) | 1.00 | (0.78-1.28) |
| Estates/Porters/Security | 1.28 | (0.95-1.73) |
| Other - no patient contact | 0.80 | (0.66-0.97) |
| Other - patient contact | 0.89 | (0.61-1.28) |
